# Amount and certainty of evidence in Cochrane systematic reviews of interventions: a large-scale meta-research study

**DOI:** 10.64898/2025.12.19.25342674

**Authors:** Thomas Starck, Philippe Ravaud, Isabelle Boutron

## Abstract

**Background:** Despite continuous growth in primary research production, it remains unclear whether this translates into more eligible studies for systematic reviews and improved evidence certainty.

**Objectives:** To quantify the amount and certainty of evidence in Cochrane systematic reviews of interventions and describe its evolution.

**Design:** Large-scale meta-research study.

**Data source:** Cochrane Database of Systematic Reviews (April 8, 2025).

**Eligibility criteria:** Cochrane intervention reviews reporting “Summary of findings” tables.

**Data extraction:** Data were extracted using web scraping and a validated large language model.

**Analysis:** We described evidence amount (included studies and participants) and GRADE certainty for each population-intervention-comparison-outcome (PICO) question in “Summary of findings” tables, comparing initial versions with latest updates.

**Results:** Among 5,116 included reviews (64,849 PICOs), 24% (n=15,768) found no eligible studies, 31% (n=20,390) had 1 study, 14% (n=8,796) had 2, and 31% (n=19,895) had >2. Included studies were 97% randomized trials. Median [Q1-Q3] participant count was 123 [0–557]. Certainty was high for 4% (n=2,852), moderate for 16% (n = 10,574),, and low/very low/unassessed for 80% (n = 51,423). There was no improvement over time. Of 7,461 updated PICOs (median [Q1-Q3] interval 4.3 years [2.6-6.4]), the number of included studies remained identical for 63%; certainty was unchanged for 71%, upgraded for 13%, and downgraded for 15%.

**Limitations:** Certainty changes across updates may reflect evolving assessment practices or error correction rather than genuine evidence changes.

**Conclusion:** The amount of evidence in Cochrane reviews barely changed over 15 years, with no improvement in certainty.

**Implications of key findings:** To increase evidence quantity and quality, our findings suggest the need to direct primary research toward knowledge gaps and reduce avoidable trial waste.

**Primary Funding Source:** None.

## INTRODUCTION

Clinical research generates an ever-increasing volume of scientific knowledge, with an estimated 35,000 new reports of clinical trials indexed in PubMed in 2020 alone [1]. Given this information overload, rigorous evidence syntheses, such as Cochrane reviews, are essential for decision-making. The Cochrane collaboration has published more than 9,000 reviews, with 66% of new World Health Organization guidelines in the past five years citing at least one of their reviews [2].

However, despite this massive growth in primary research production, confidence in systematic reviews remains limited by the actual availability and quality of evidence for specific clinical questions [3–6]. To communicate how much confidence decision-makers can place in these syntheses, the Grading of Recommendations Assessment, Development and Evaluation (GRADE) framework has become the reference standard [7]. First presented in 2004 [8] and adopted by Cochrane in 2008 [9,10], GRADE evaluation has been progressively incorporated into Cochrane systematic reviews, becoming mandatory in the mid-2010s as part of a “Summary of findings” table [11].

Previous studies have documented low certainty of evidence in samples of Cochrane reviews across multiple clinical fields [12–20], yet the amount of primary evidence underlying each individual PICO question, and whether it has grown over successive review updates, has never been systematically quantified.

This study aimed to quantify the amount (i.e., number of included studies and participants) and certainty of evidence in Cochrane systematic reviews of interventions, and to describe how this has evolved over time.

## METHODS

This meta-research study was reported in accordance with the guidelines for reporting meta-epidemiological methodology research [21]. No protocol was pre-registered for this study.

### Eligibility Criteria

We included all Cochrane systematic reviews of interventions (i.e., reviews assessing the effectiveness and/or safety of a treatment, vaccine, device, preventive measure, procedure or policy) that reported a “Summary of findings” table. We excluded other types of reviews such as diagnostic test accuracy reviews, prognosis study reviews, qualitative evidence syntheses, methodology reviews, overviews of reviews, rapid reviews and prototype reviews. We also excluded network meta-analyses.

### Search strategy

We developed an R-based pipeline to automatically scrape all the versions (initial version and subsequent updates where available) of all the systematic reviews indexed in the Cochrane Database of Systematic Reviews [22], their associated metadata and, where available, their “Summary of findings” tables (Figure 1, Supplementary Figures S11 and S12). The scraping was launched on April 8, 2025.

**Figure 1.**
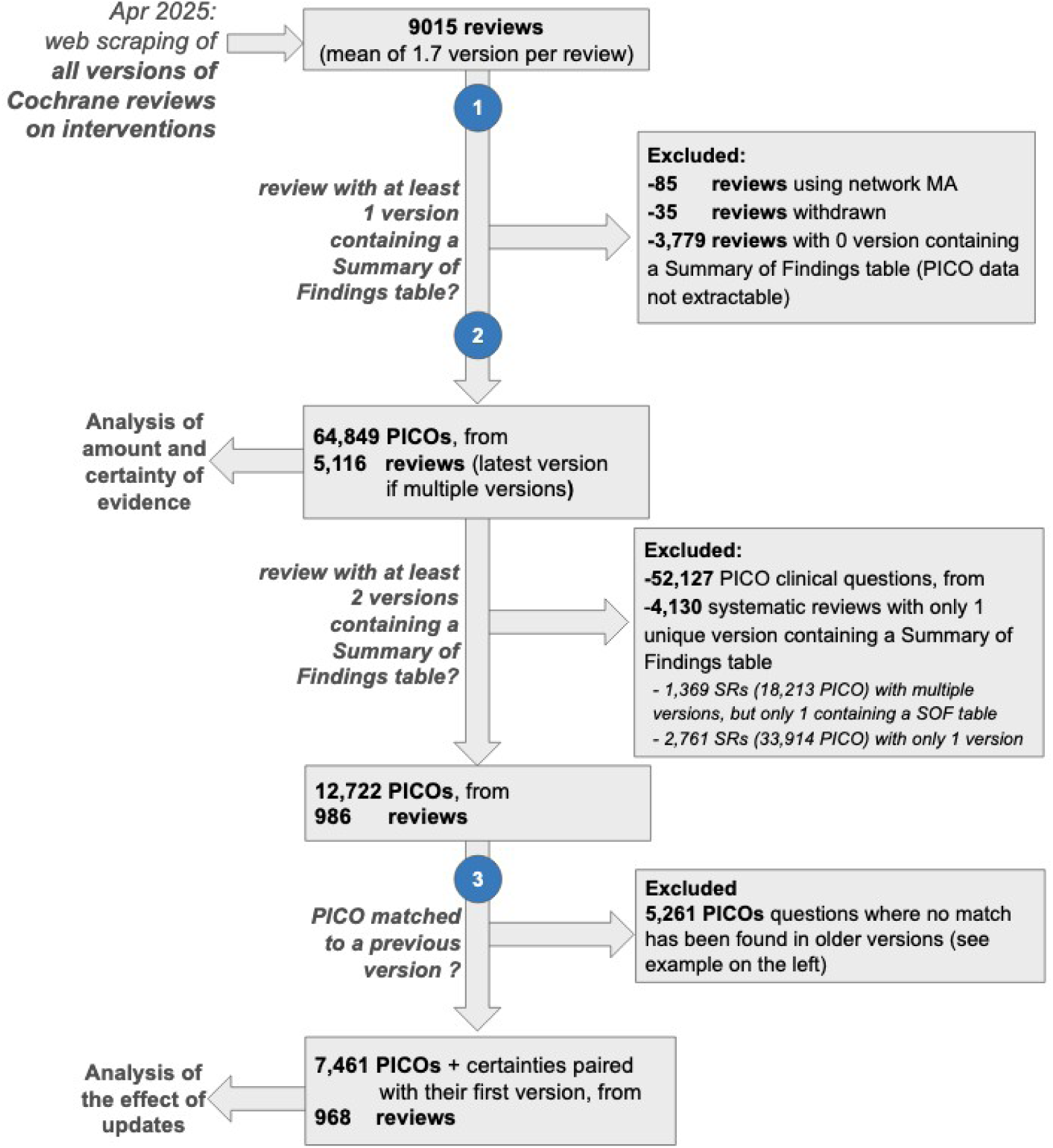
Flow chart of the included PICO questions (combinations of population-intervention-comparison-outcome). Numbers in blue disks indicate an LLM-assisted task. 1: extraction of the review literature search dates from abstracts; 2: extraction of the “Summary of findings” table contents (PICO elements, certainty of evidence, number of included participants, number of included studies). 3: matching of PICOs in the most recent review update to PICOs in previous review versions.

### Data retrieved through web scraping

For each review version, the following data were automatically extracted during our web scraping:

- metadata: identifier, version, title, URL
- ”Search Methods” text from the abstracts: this was then processed by a large language model (LLM) to extract the literature search dates.

### Data extraction assisted by a large language model (LLM)

We used LLMs to extract search dates and Summary of Findings tables data, and match PICO questions across review versions, when several versions of a review existed. The specific procedures, including prompt optimization and error definitions, are detailed in the Supplementary Materials (Sections B and C), and Supplementary Methods.

### Search date extraction

Because the publication date of the reviews can differ substantially from the actual search date (Supplementary Figures S13 and S14), we extracted the search month and year from the “Search Methods” section of each abstract using GPT-o4-mini-high. Validation on an independent testing set of 600 search dates manually extracted by two trained reviewers showed that the LLM exactly matched the human extraction in 84% of cases; 98% were within ±1 year.

### ”Summary of findings” tables extraction

We used GPT-o3-mini-high with zero-shot prompting on the raw HTML code displaying the Summary of Findings tables to extract: PICO elements, number of included studies (including whether they were randomized) and participants, and the certainty of evidence for each outcome (see Supplementary Figure S12 for the structure of a “Summary of findings” table on the Cochrane website).

Two specifically trained medical students manually assessed the extraction of a random sample of 2,660 PICO questions from 92 unique reviews. Overall, 98% of PICO questions were correctly extracted. Only 1% were missed and 1% contained major altering errors (dropping to 0.2% when a single highly problematic review was excluded). Inter-reviewer agreement on a duplicate subset of 25 reviews (309 PICOs) was 99-100% (we did not use Cohen’s kappa to judge agreement because the error rates were very low [23]).

### Automatic matching of similar PICO questions with previous versions of the review

To assess the evolution of evidence, we matched similar PICO questions present in the most recent update of a review and its initial versions that reported a “Summary of findings” table. Because the wording of PICO elements can vary across versions (e.g., “participants with” vs “people with”), we used GPT-o4-mini-high with few-shot prompting (Supplementary Section C.3).

We provided the LLM with an Excel sheet containing the review version identifiers and the previously extracted PICO elements. A PICO question was classified as similar between review versions only if all of its elements (population, intervention, comparison, and outcome) were judged similar; questions without a clear equivalent were left unmatched.

For quality control, two reviewers independently evaluated a random sample of 4,853 PICO questions from 181 reviews. Overall, the human reviewers agreed with the LLM’s matching decisions in 98% of cases (inter-reviewer reliability: Cohen’s κ=86%). A more granular assessment of the similarity between matches showed complete agreement (82%), mostly agree (16%), mostly disagree (1%), and complete disagreement (0.6%). Further details on the agreement for each individual PICO element are available in Supplementary Section B.4.

### Analyses

Given the low error rates of the LLM-assisted tasks, the analysis was carried out on the entire raw database.

First, we described the amount (number of included studies and participants) and certainty (high, moderate, low, very low, unassessed) of evidence across all Cochrane reviews. If a review had multiple versions, we included only the most recent update.

Second, for reviews with at least two versions that reported a “Summary of findings” table, we described the evolution of the evidence between the initial and latest version.

### Role of the funding source

No funding was required for this study.

## RESULTS

### Identification and characteristics of Cochrane Systematic Reviews

The flow chart is reported in Figure 1.

We identified 9,015 systematic reviews on interventions. Overall, 56% (n = 5,022) were not updated (i.e., one version), 27% (n = 2,416) had one update (i.e., two versions); 11% (n = 978) had two updates (i.e., three versions), and 7% (n = 599) had three or more updates. The median [Q1–Q3] delay to update was 4.5 [2.9–6.8] years.

We identified 5,116 reviews with at least one version reporting a “Summary of findings” table (Supplementary Figure S1), yielding a total of 64,849 PICO questions; for reviews with multiple versions, only the most recent version was analyzed. On average, these reviews had 2.3 “Summary of findings” tables. The median number of PICO questions assessed per review was 8 [Q1–Q3: 6–16].

### Evidence available for each PICO question

#### Amount of eligible evidence

Of the 64,849 PICO questions, 24% (n = 15,768) had no studies reporting the outcome; 31% (n = 20,390) had only one study included, 14% (n = 8,796) had two studies, and 31% (n = 19,895) had three or more studies (Table 1). Half (50%, n = 2,575) of the reviews had at least one PICO for which no study was identified. Nearly all PICOs (97%) with at least one included study relied exclusively on RCTs. The median [Q1–Q3] number of included participants was 123 [0–557].

**Table 1.** Evidence included (number of studies and participants) in assessments of the certainty of evidence for PICO questions in Cochrane reviews, and evolution of the included evidence when reviews were updated.

| **Evidence included in assessments of the certainty of evidence for PICO questions.**<br>64,849 PICO questions from 5,116 Cochrane reviews with a “Summary of findings” table<br>(only the most recent review update was analyzed if multiple versions existed). |  |  |  |
| --- | --- | --- | --- |
| Number of included studies | % of PICO questions | Number of included participants | % of PICO questions |
| 0 | 24% | 0 | 25% |
| 1 | 31% | 1–100 | 21% |
| 2 | 14% | 101–500 | 27% |
| >2 | 31% | >500 | 27% |
| **Evolution of included evidence after a review update (change from the first to the most recent review version)**<br>7,461 PICO questions from 986 Cochrane reviews |  |  |  |
| Number of included studies | % of PICO questions | Number of included participants | % of PICO questions |
| identical | 63% | identical | 59% |
| increase | 33% | increase | 35% |
| decrease | 4% | decrease | 6% |

The amount of evidence included per PICO in reviews was stable over time (Figures 2 and Supplementary Figure S2).

**Figure 2.**
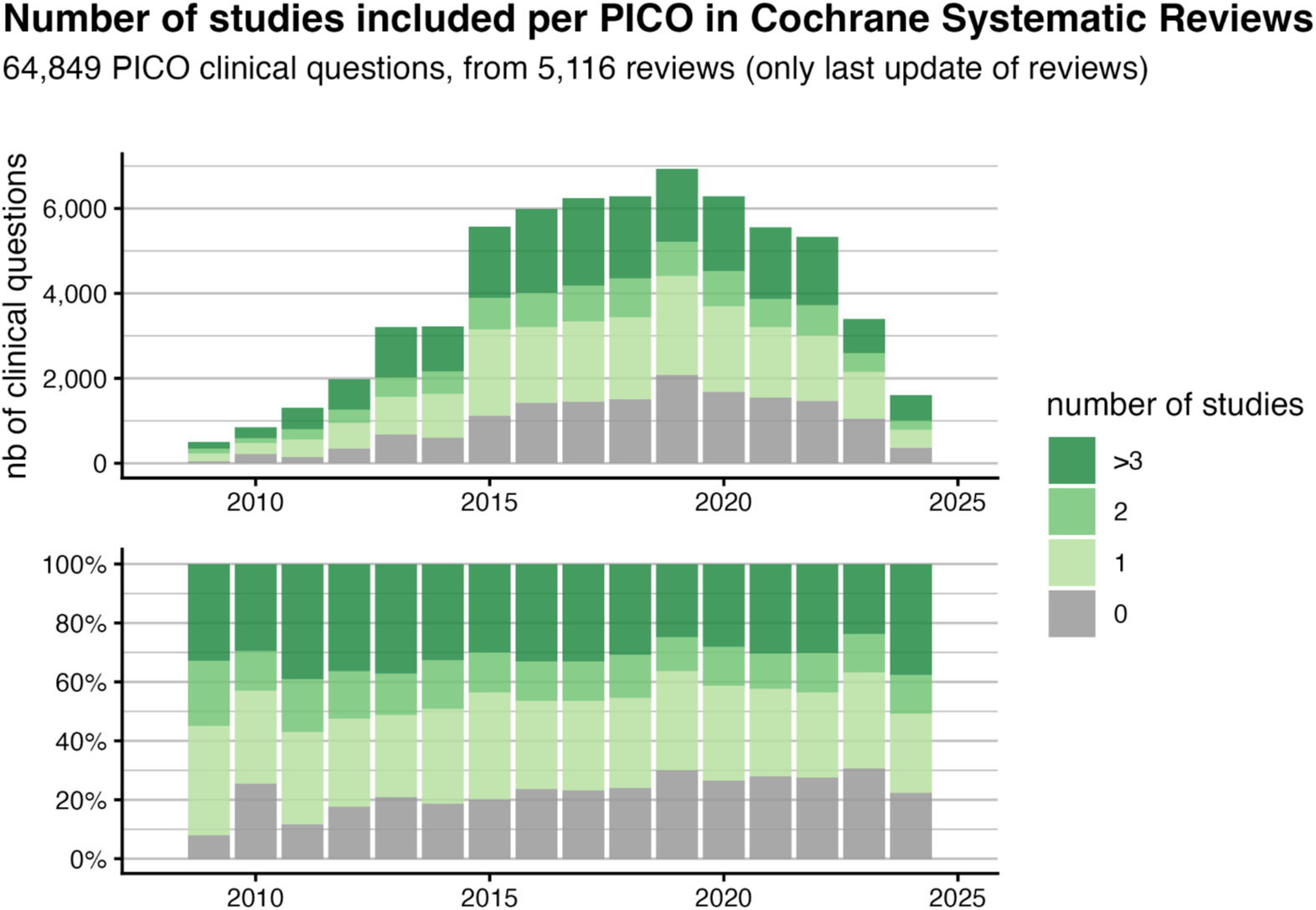
Top: number of PICO questions assessed each year in Cochrane reviews with a “Summary of findings” table, with the number of studies included for the PICOs. Bottom: relative share of each category of number of included studies, for each year.

### Certainty of evidence

#### At the PICO question level

Among the 64,849 PICO questions reported in the “Summary of findings” tables, the certainty of evidence was rated as high for 4 % (n = 2,852), moderate for 16% (n = 10,574), low for 27% (n = 17,409), very low for 26% (n = 17,012), and not assessed for 26% (n = 17,002) because o f insufficient data.

Over the 15-year period studied, there was no overall improvement in the certainty of evidence (Figure 3). Supplementary Figures S3 and S4 show the certainty of evidence according to the detailed distribution of the number of included studies (Supplementary Figure S3) and participants (Supplementary Figure S4) per PICO, with the PICO-associated certainty.

**Figure 3:**
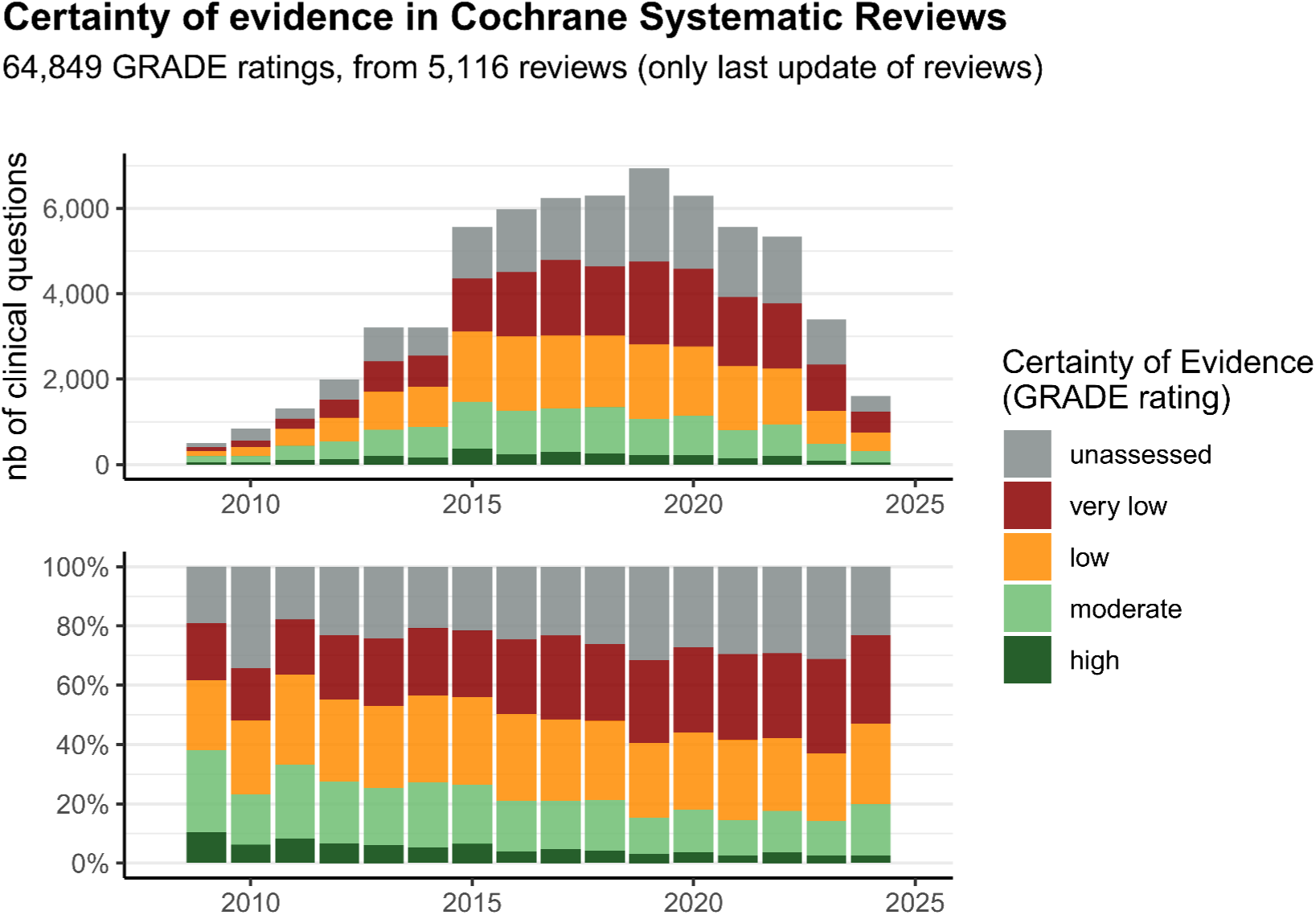
Top: number of PICO questions assessed each year in Cochrane reviews with a “Summary of findings” table, for the different certainties of evidence. Bottom: relative share of each certainty of evidence, for each year.

### At the review level

Of the 5,116 systematic reviews with a “Summary of findings” table, 17% (n = 884) included at least one PICO question for which the evidence was rated as high certainty and 55% (n = 2,829) included at least one PICO question for which the evidence was rated as moderate or high. In contrast, 45% (n = 2,287) of reviews included only PICO questions for which the evidence was rated as low, very low, or unassessed. Only 5% (n = 269) of reviews included only PICO questions with evidence exclusively rated as “moderate” or “high”.

#### Evolution of evidence after an update Amount of eligible evidence

Of the 64,849 PICOs (5,116 reviews), 52,127 PICOs (4,130 reviews) could not be matched to a previous assessment; 33,914 (2,761 reviews) had a unique version, and 18,213 (1,369 reviews) had several versions but only one version reported a “Summary of findings” table.

In addition, among the reviews that were ultimately included, 5, 261 PICOs could not be matched to a previous assessment.

This left 7,461 PICOs (986 reviews) included in the analysis for the effect of updates. The number of studies included in the most recent update remained identical to the number included in the original version for 63% (n = 4,721) of PICO questions, increased for 33% (n = 2,426), and decreased for 4% (n = 314) (Table 1). For PICOs with an increase in the number of included studies, the median [Q1–Q3] number of additional studies was 2 [1–4]. For PICOs with an increase in the number of included participants, the median [Q1–Q3] number of additional participants was 315 [117–1,073].

Increases in the number of included studies were more frequent when the interval between the first and last review version was longer (Figure 5). When the interval was shorter than 5 years, the number of included studies was unchanged for 72% of PICOs (n=3,361), increased for 25% (n=1,181), and decreased for 3% (n=132); when it exceeded 5 years, 49% (n=1,360) remained unchanged, 45% (n=1,245) increased, and 7% (n=182) decreased. Decreases in the number of included studies may reflect stricter eligibility criteria in updated reviews, and in some cases extraction errors.

**Figure 4:**
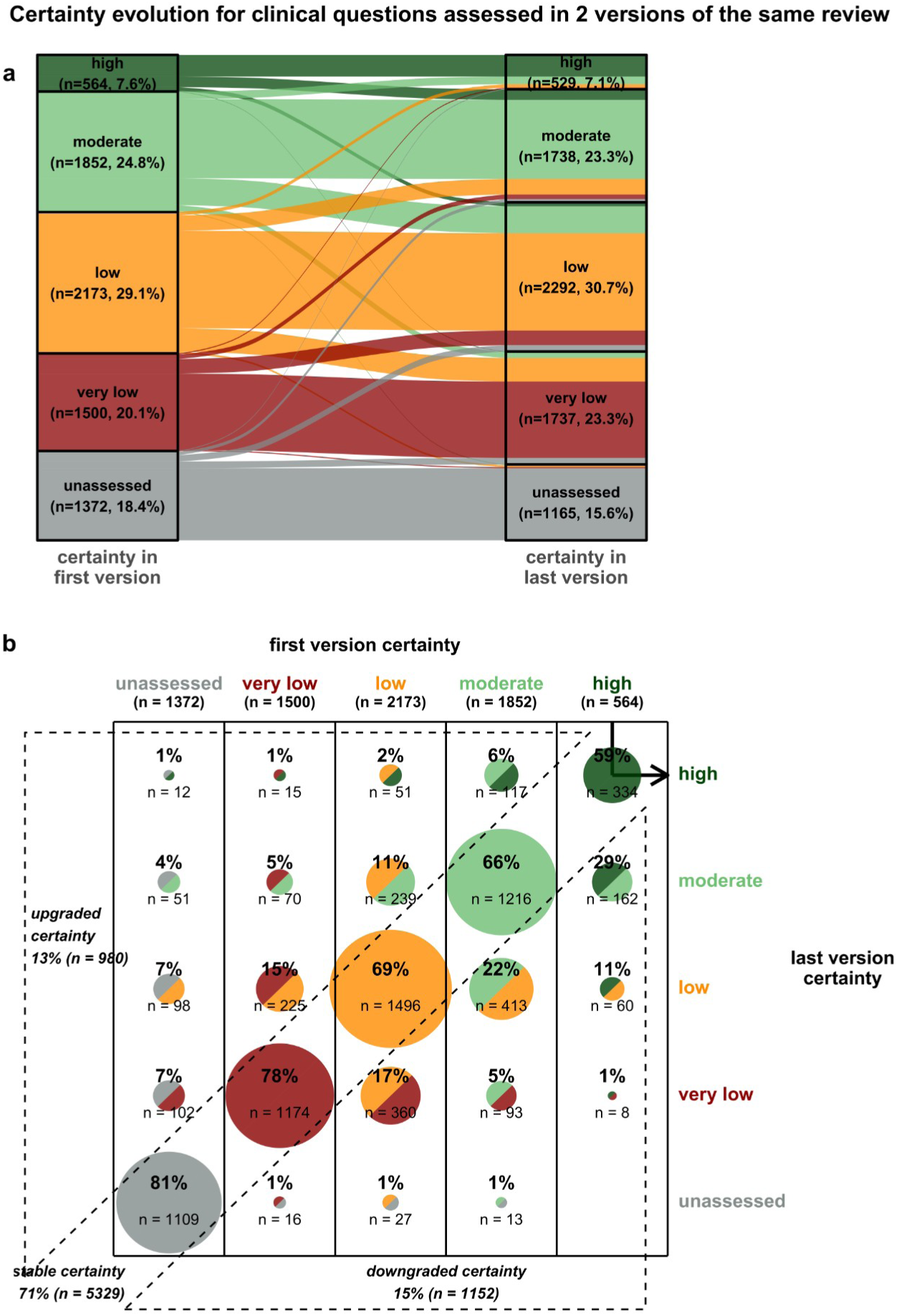
Evolution of the certainty of evidence for the 7,461 pairs of PICO questions (assessed in 986 reviews) for which more than one review version was available. **Top**: Sankey diagram displaying the evolution from the first to the most recent version. **Bottom**: full table of transitions between the first and the most recent version. As an example of how to read this table, the information indicated by the arrow in the top right corner would be interpreted as follows: *out of the 564 certainties graded high in the first review version, 334 (59%) remained high in the most recent version of the same review.* On the diagonal are unchanged certainties; in the bottom right, downgraded certainties; and in the top left, upgraded certainties.

**Figure 5:**
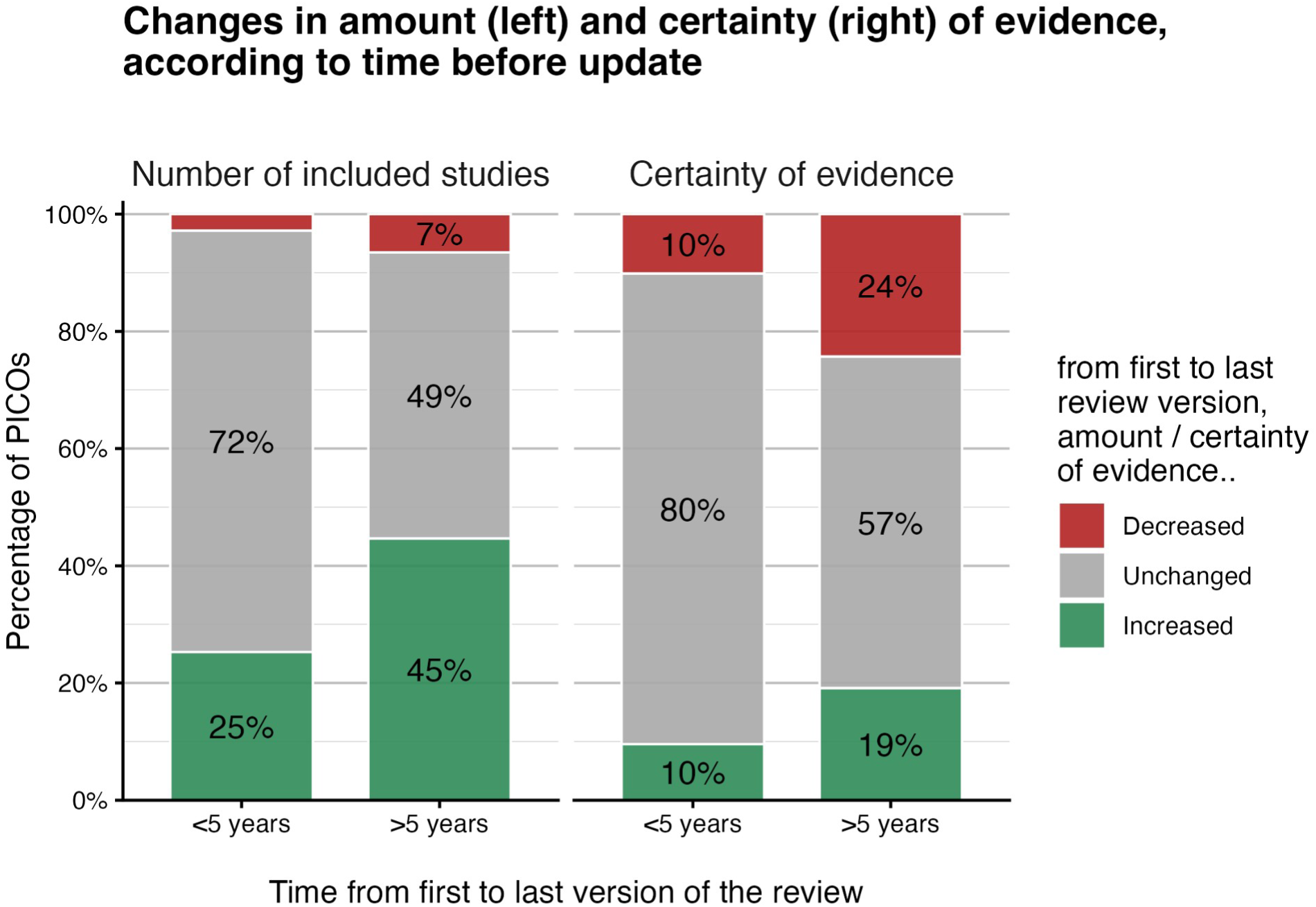
Subgroup analysis according to time to update (<5 years vs >5 years). Stacked bars show the proportion of matched PICO questions for which the number of included studies (left panel) and the certainty of evidence (right panel) increased, remained unchanged, or decreased between the first and last review version.

#### Certainty of evidence

Figure 4 displays the evolution in the PICO certainty of evidence between the initial review version and the most recent update. Overall, the certainty of evidence did not improve: the certainty of evidence for 8% (n = 564) of PICOs in the initial versions of the reviews was rated as high, compared with 7% (n = 529) in the most recent updates. The transition table in Figure 4b highlights that out of the initial 7,461 assessed certainties, 71% (n = 5,329) remained similar; 13% (n = 980) were upgraded to a higher certainty, while 15% (n = 1,152) were downgraded to a lower certainty. Extreme transitions (e.g., from high to very low) were very uncommon.

Figure 5 shows that changes in certainty of evidence also depended on time to update. Longer intervals were associated with more frequent changes in certainty, without a marked shift towards improvement. When the interval between the first and last review version was shorter than 5 years, certainty of evidence remained unchanged for 80% (n=3,753) of PICOs, while 10% (n=447) were upgraded and 10% (n=474) downgraded. When the interval exceeded 5 years, the proportion unchanged fell to 57% (n=1,576), while 19% (n=553) of PICOs were upgraded and 24% (n=678) downgraded.

### Sensitivity analysis

To explore why the certainty of evidence was sometimes downgraded, we performed a post-hoc subgroup analysis restricted to PICOs for which no new evidence was identified (Supplementary Figure S10). When no new evidence was included, certainty was downgraded in 10% of updated PICOs and upgraded in 4%. Moreover, the longer the interval before an update, the higher the probability of downgrading, increasing from <5% within the first two years to 20–30% after seven years. Since no additional evidence was incorporated, these downgrades probably reflect changes over time in how certainty of evidence is assessed.

## DISCUSSION

### Statement of principal findings

We analyzed the evidence synthesized in 5,116 Cochrane reviews reporting a “Summary of findings” table for a total of 64,849 PICO questions. Overall, most PICO questions relied on little evidence: 25% included no studies, 68% included 2 studies or fewer, and 46% included 100 participants or fewer. Consequently, the certainty of evidence was most often low or very low, with high certainty very uncommon (i.e., 4% of PICOs). Updating a review seldom changed the amount of eligible evidence: even after more than 5 years, for about half of the PICOs no additional studies were included.

### Strengths and weaknesses of the study Strengths

To our knowledge, this is the first study to describe the certainty of evidence and its evolution on such a large sample. We included all Cochrane intervention reviews, a large and methodologically rigorous corpus. They include a wide diversity of PICO questions, and apply a process to select research questions and outcomes considered most relevant to clinical practice and public health.

### Limitations

This study has several limitations. First, data were extracted exclusively from the “Summary of findings” tables of systematic reviews of interventions published in the Cochrane Library, which can limit the generalizability of the results. Further, data extraction was automatically performed using an LLM, which may have introduced classification or extraction errors; however, a manual quality control process was applied and confirmed high accuracy rates, which supports the robustness of the extracted data.

In addition, we must recognize that Cochrane reviews are not immune to error, and meta-research has documented inconsistencies in the application of GRADE within Cochrane reviews [23]. Some of the certainty changes we observe may therefore reflect error correction rather than genuine changes in the evidence base. Likewise, during the studied period, there was a change from the original Risk of Bias [24] tool to RoB 2 [25]. These factors might explain that even when no evidence is identified, the certainty of evidence was downgraded in 10% of updated PICOs and upgraded in 4%. However, this limitation only concerns the assessment of certainty of evidence, but not the objective description of the amount of eligible evidence.

Another limitation is that our analysis was restricted to pairwise systematic reviews. We excluded network meta-analyses (NMAs), which represented less than 2% of the eligible reviews (n=85). Their “Summary of findings” tables are displayed differently, making automatic extraction overly complex for a minimal marginal gain. Moreover, the GRADE framework applied to NMAs (e.g., the CINeMA approach) evaluates mixed evidence nodes and includes additional domains not applicable to pairwise meta-analyses, which would have introduced heterogeneity in our certainty assessments.

### Strengths and weaknesses in relation to other studies, discussing important differences in results

To our knowledge, this is the first study to systematically assess the amount of evidence associated with each PICO in Cochrane Summary of Findings tables. Previous studies that quantified the number of studies and participants were restricted to the review level [26] and did not assess evidence at the outcome level.

Our results are consistent with previous cross-sectional studies assessing the certainty of evidence in smaller samples of Cochrane reviews [12–16,27], or restricted to specific medical fields [14,17–20,28]. We identified only two studies assessing the evolution of the certainty after an update on a limited sample of Cochrane reviews. They also found that most updates (50 to 70%) did not change the certainty of evidence [15,16].

Finally, our results are in line with the results of studies evaluating the level of evidence of clinical practice guidelines recommendations for various organizations in different fields [29–41]. Overall, only 4% to 30% of clinical practice guideline are supported by a high level of evidence.

### Meaning of the study: possible explanations and implications for clinicians and policymakers

Despite the continuous growth in primary research production, with over 35,000 new reports of clinical trials indexed in PubMed for 2020 alone [1], most clinically relevant PICO questions identified in Cochrane systematic reviews remain informed by very few studies, and this scarcity has not improved over 15 years of review updates (Figure 2). One quarter of all PICO questions had no eligible study, and two thirds were informed by two studies or fewer (Table 1). Even after more than five years, half of the PICOs did not include new studies; and among those that did, this did not translate into a clear improvement in certainty of evidence (Figure 5, Supplementary Figure S10). This pattern suggests that the problem is not only one of scarcity of evidence, but also of the methodological limitations and informativeness of the studies being produced.

Our results question the efficiency of the current evidence ecosystem, which is unable to adequately support decision making [6]. There is a need to bridge the gap between evidence generation, evidence synthesis and guidelines development to develop a learning system [5,42]. We point to three concrete axes for improvement.

First, primary research could be directed toward questions where evidence is most lacking. The 64,849 PICO questions from the Cochrane Library with their associated certainty levels documented here constitute an empirical, large-scale evidence gap map. Funders, investigators, and policymakers could systematically use this map to prioritize research questions and align outcome reporting with existing evidence gaps. Such a feedback loop between evidence synthesis and evidence generation is currently absent from most research governance frameworks.

A second axis is to reduce the avoidable research waste in the conduct and reporting of RCTs [43]. It is estimated that 42% of research waste in trials is due to inadequate methods (inadequate allocation concealment and sequence generation, absence of blinding of outcome assessors, and incomplete outcome data) that could be partly avoided by simple and inexpensive adjustments [3]. For incomplete outcome reporting, Yordanov et al. [42] showed that 78% of trials included in a Cochrane review were excluded from at least one meta-analysis because key outcomes were not appropriately planned or incompletely reported. Reducing this waste, through better coordination of trial design, prospective outcome harmonization, and stricter reporting standards, would allow the existing volume of RCTs to contribute more effectively to the evidence base.

Third, while Cochrane reviews almost exclusively rely on RCTs, considered as the gold standard of therapeutic evaluation, there are clinical questions where RCT evidence is structurally unlikely to accumulate and for which alternative sources of evidence could be considered. RCTs are costly (tens to hundreds of millions of US dollars) [44], raise major recruitment challenges (from 1997 to 2020, 4 out of 10 NIHR-funded RCTs did not meet their recruitment target sample size) [45], and can have limited external validity. For a significant subset of PICO questions, including rare diseases, rare adverse events, the low number of eligible studies and included participants documented here suggests that RCT evidence will struggle to reach the volume required to achieve high certainty. For these questions, the increasing availability of high-quality routinely collected data [46] and novel frameworks such as target trial emulation [47] offer promising complements to randomized evidence, with the potential to provide faster and broader evidence. However, this non-randomized evidence comes with its own sources of bias, data quality challenges, and risks of introducing additional heterogeneity into evidence syntheses. Realizing its potential will therefore require the development of robust methodological standards for integrating such evidence into systematic reviews and GRADE assessments, a necessary safeguard before this approach can be broadly applied.

### Unanswered questions and future research

Future work should investigate concrete mechanisms to establish feedback loops between evidence synthesis and primary research generation, ensuring that the specific evidence gaps identified in systematic reviews directly inform funders on which research questions to prioritize. A key outcome would be whether such feedback loops increase the volume and certainty of evidence over subsequent years. Additionally, establishing robust methods for incorporating high-quality non-randomized evidence into decision-making frameworks remains a critical methodological priority.

## CONCLUSION

In this large-scale assessment of Cochrane systematic reviews of interventions, we found that the amount of available evidence was frequently limited: one quarter of all PICO questions had no included studies, and two thirds were informed by either one or two studies. High-certainty evidence was uncommon, and most certainties were low, very low, or unassessed. Updates rarely brought new eligible evidence for most PICOs, and consequently certainty usually did not improve.

These findings reveal a structural misalignment between primary research production and critical evidence gaps, and point to three axes for improvement: directing primary research toward identified knowledge gaps, through a systematic feedback loop between evidence synthesis and primary research prioritization; reducing avoidable waste in RCT conduct and reporting o reduce risk of bias and increase certainty; and developing robust methods to integrate high-quality non-randomized evidence when randomized trials are unfeasible.

## Statements

### Public and Patient Involvement statement

No patients or members of the public were involved in this research.

### Ethics approval

Ethics approval was not required according to French regulations as this study did not involve human participants, their data, or animals.

### Scraping and copyrighted content

This work complies with the terms and conditions of Wiley, the publisher of the Cochrane Library (see https://www.wiley.com/en-gb/terms-of-use/ai-principles/wiley-statement-illegal-scraping-ai-copyright). We used information from Cochrane review abstracts (publicly available) and “Summary of findings” tables (public or licensed access). No copyrighted content was reproduced or shared; only aggregate statistics are reported. No AI training or commercial reuse was involved.

### Author contributorship

TS and IB conceived the study. TS carried out the data extraction, coding, and analyses. TS, IB, and PR contributed to the interpretation of the results. TS and IB wrote the first draft of the manuscript. IB and PR provided senior guidance throughout the project, and revised the manuscript. TS is the guarantor, had full access to the data, and made the final decision to submit for publication. All authors approved the final version of the manuscript. **Transparency statement**: The lead author affirms that this manuscript is an honest, accurate, and transparent account of the study being reported and that no important aspects of the study have been omitted.

### Role of the funding source

No funding was required for this study.

### Competing interest

Isabelle Boutron is director of Cochrane France and a member of the Cochrane editorial board.

### Data sharing

Data can be made available upon request after submission of a protocol which will be evaluated by authors in relation to other ongoing projects using these data.

## Supporting information

Checklist_Meta_epidemiological_Study

## Acknowledgements

We thank Victoire Perrichon and Mazal Sultan, the two medical students who manually checked the accuracy of the AI data extraction and matching of PICO questions.

## Abbreviations

LLM: large language model
AI: artificial intelligence
PICO: population, intervention, comparison, outcome
RCT: randomized controlled trial

## Notes

### Summary of Updates

Adapted to more concise format (full text <3500 words, abstract <275 words)

