## SupplementaryMaterials for "Amount and certainty of evidence in Cochrane systematic reviews of interventions: a large-scale meta-research study"

### A Additional Analyses

#### A.1 Adoption of Summary of Findings table in Cochrane reviews

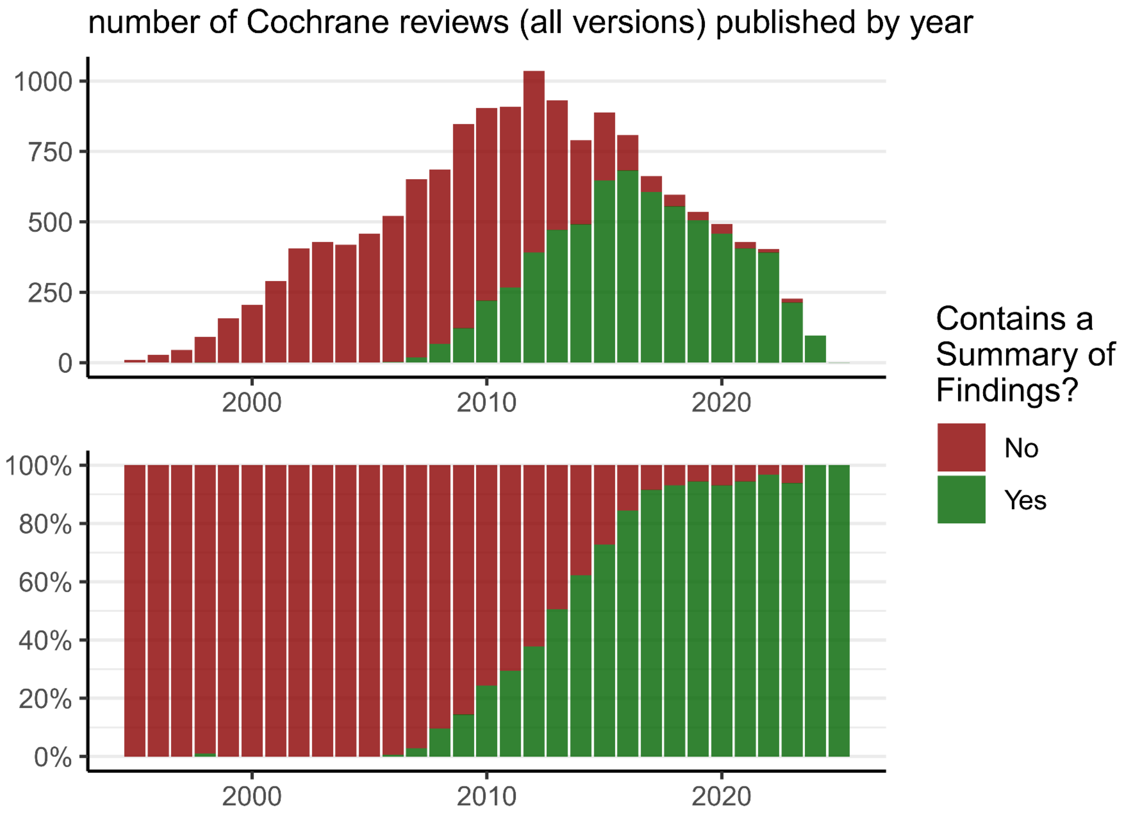


Figure S1: Presence of a Summary of Findings table in Cochrane systematic reviews through time, determined by our web scraping procedure. Note that one systematic review can be updated and thus have multiple versions. Here we present all existing versions. In this article, we only focus on reviews with a Summary of Findings table, in green.

#### A.2 Detailed information on the number of studies and participants per PICO

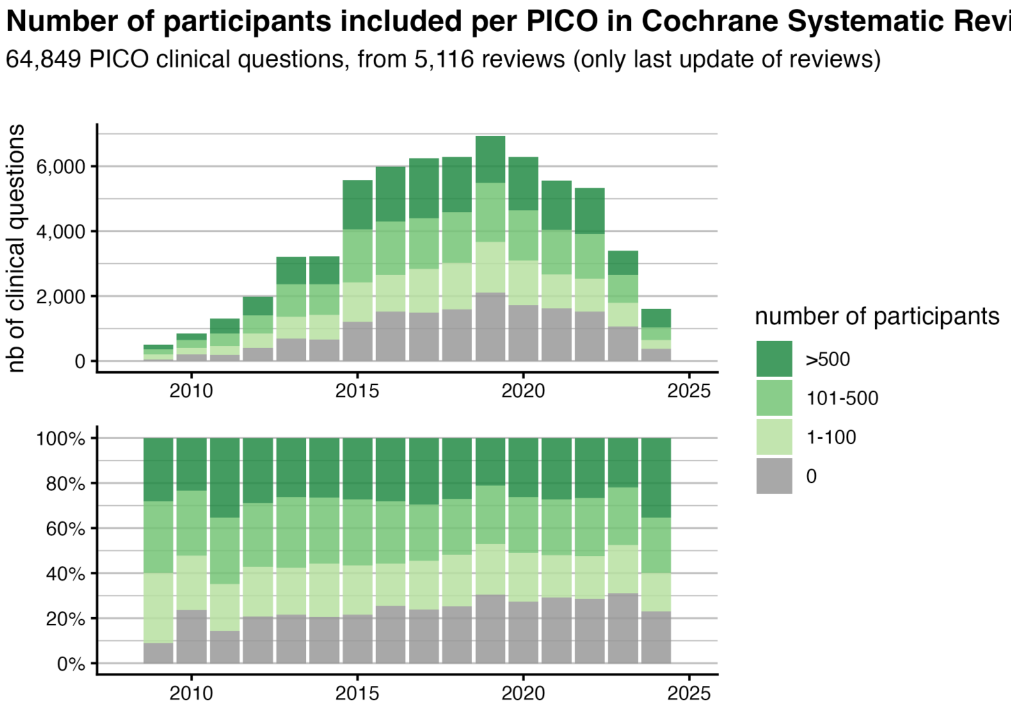


Figure S2: Temporal evolution of the number of included participants per PICO in Cochrane reviews and the literature search date of the review.

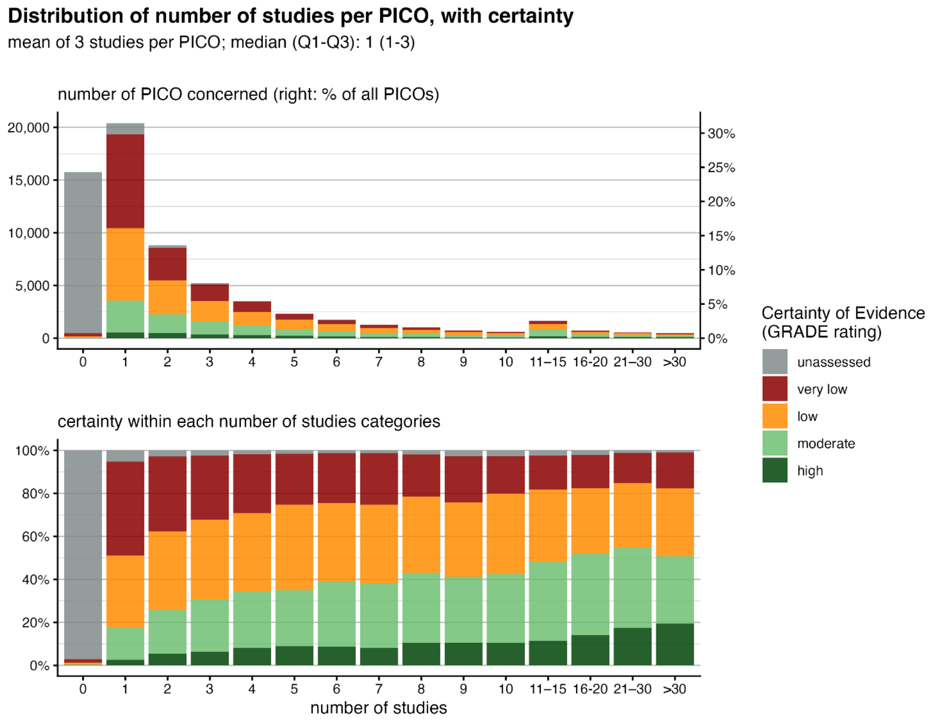


Figure S3: Distribution of the number of studies per PICO in Cochrane reviews, with the associated PICO certainty.

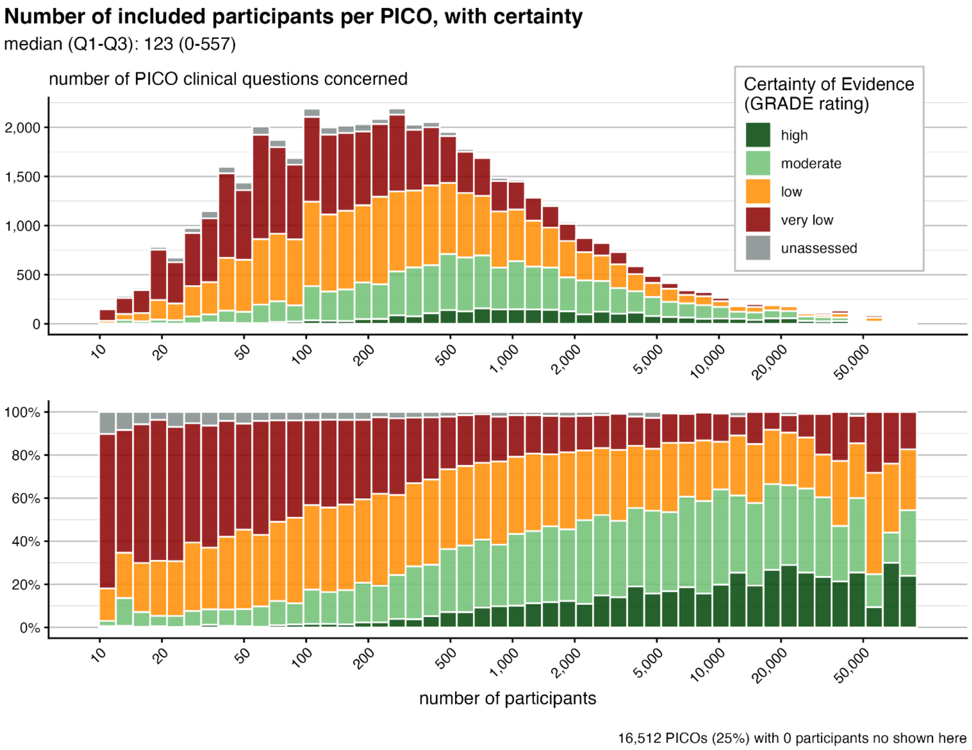


Figure S4: Distribution of the number of participants per PICO in Cochrane reviews, with the associated PICO certainty. 15,512 (25%) PICOs with 0 participants not shown here.

#### A.3 Change in the quantity of evidence after an update, depending on the time between review versions

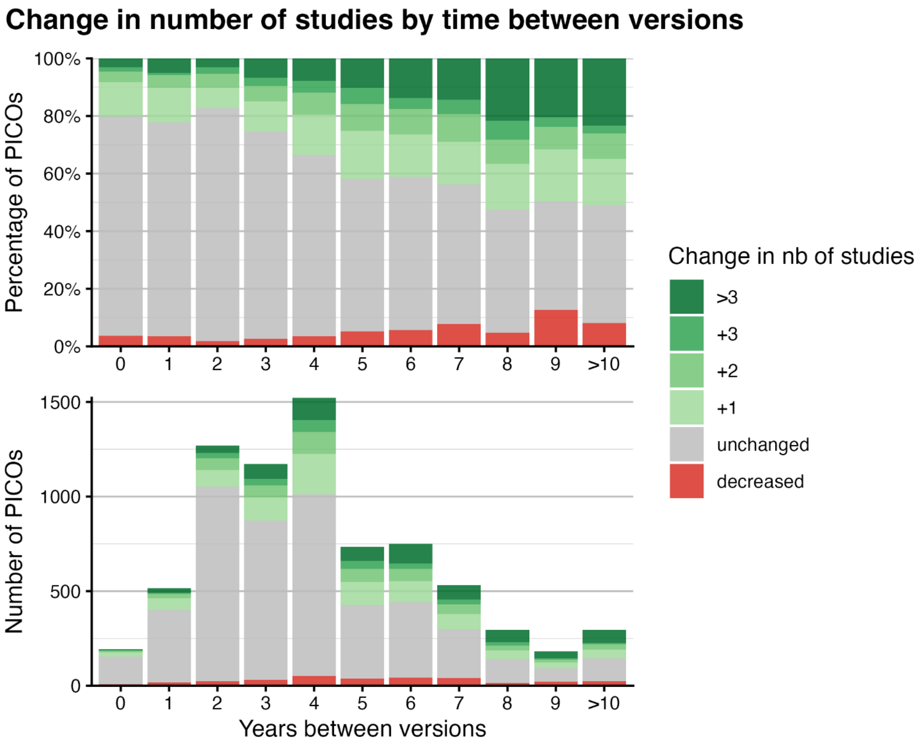


Figure S5: Change in number of included studies, depending on time before the PICO update

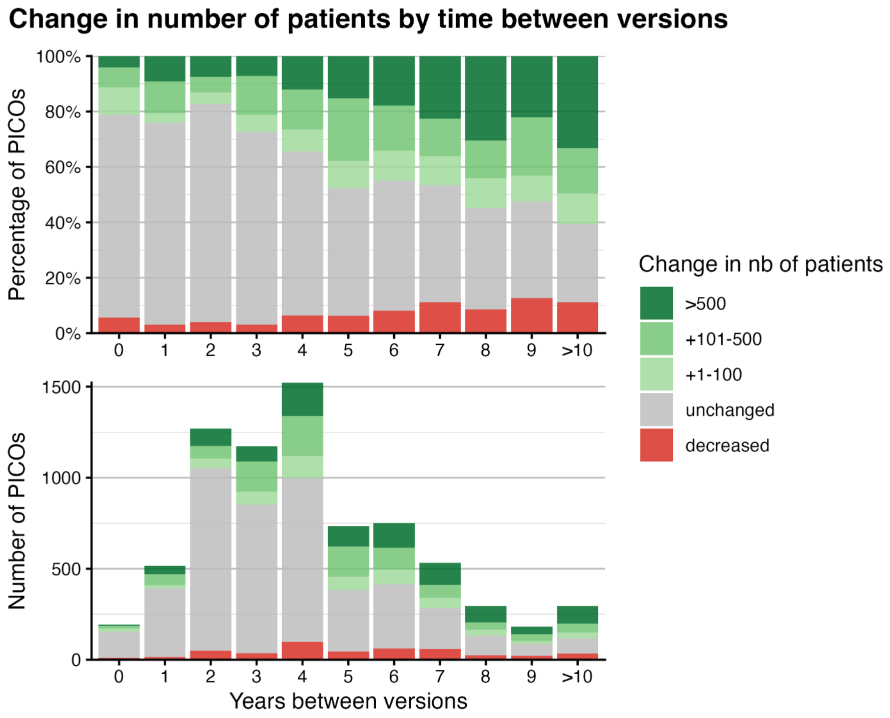


Figure S6: Change in number of included participants, depending on time before the PICO update

##### A.4 Subgroup analysis: effect of duration between first and last review versions (<5 years vs >5 years) on the certainty of evidence

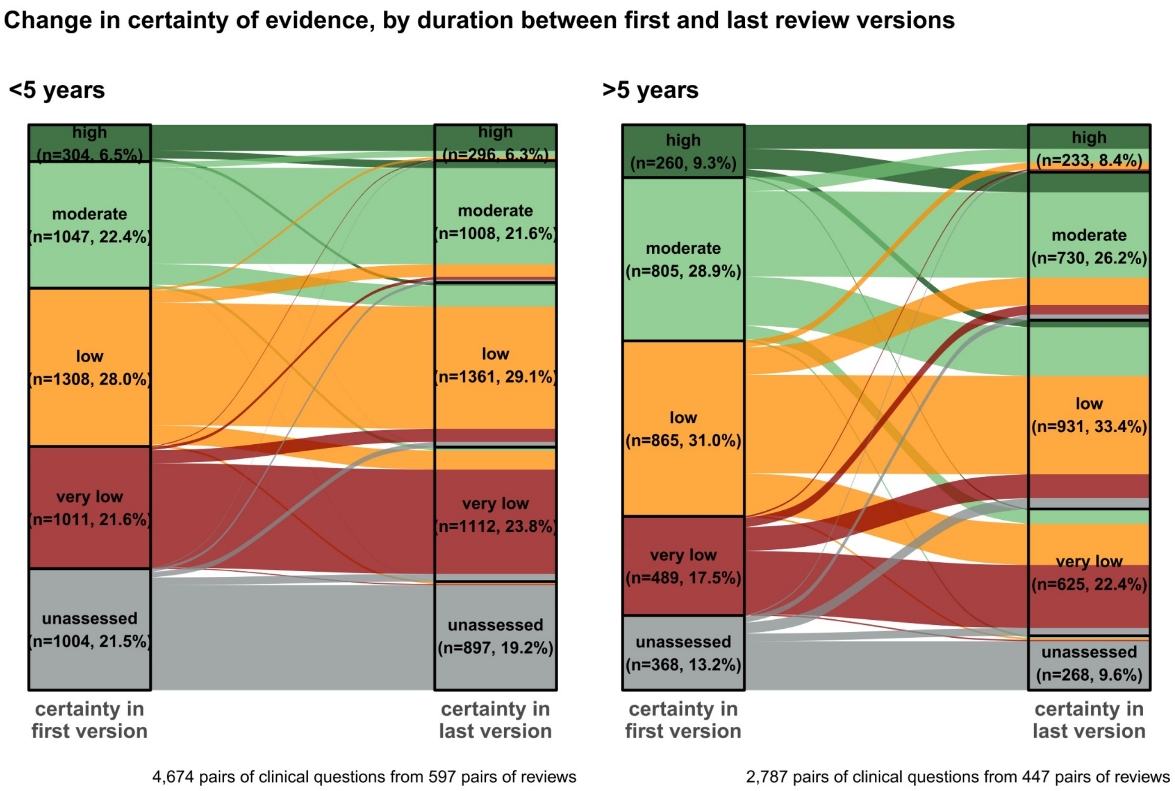


Figure S7: Subgroup analysis: sankey diagram of certainty change for reviews whose final update occurred <5 years after the first version (left) or >5 years (right).

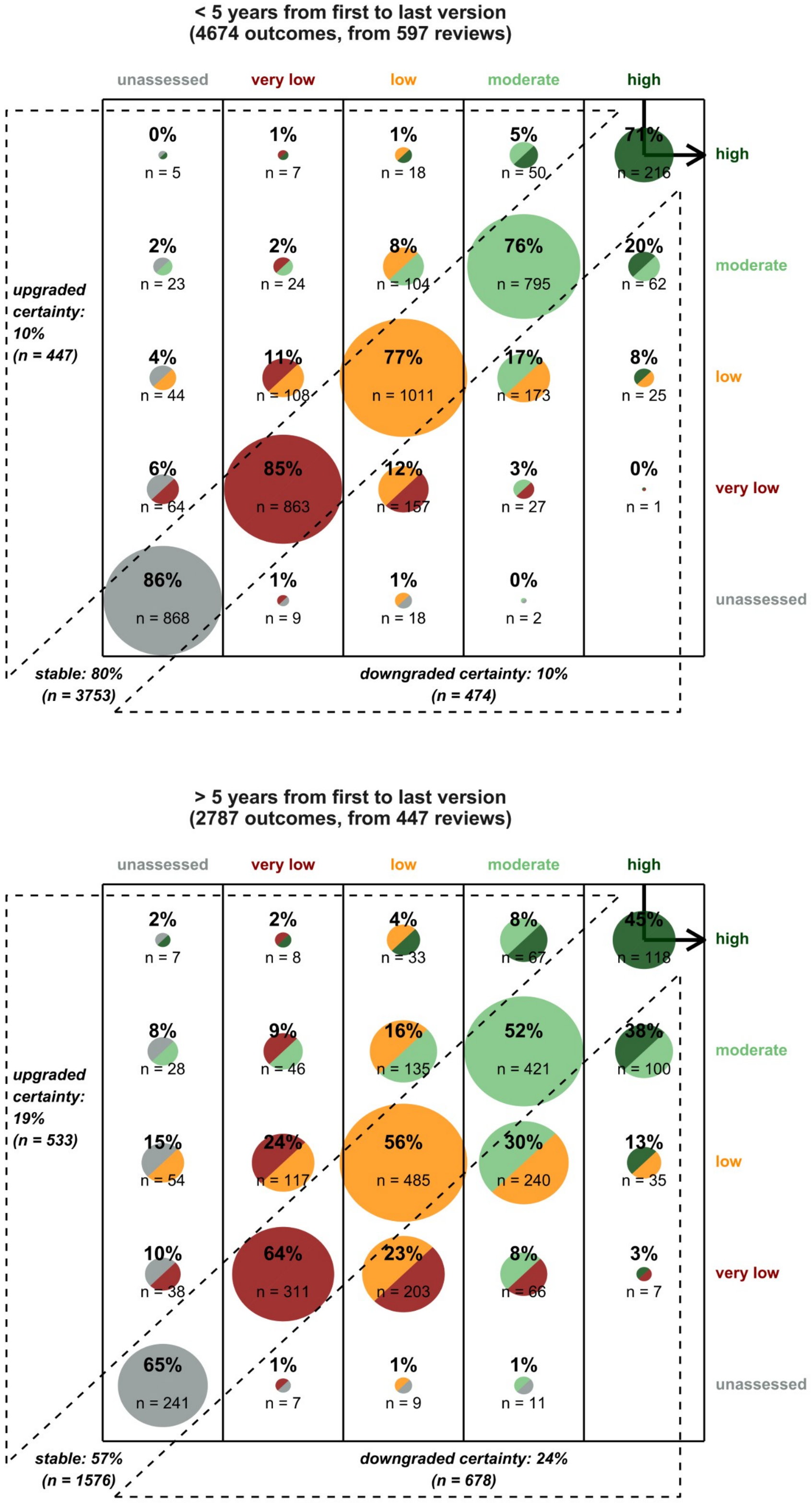


Figure S8: Subgroup analysis: full transition table for reviews whose final update occurred <5 years after the first version (top) or >5 years after the first version (bottom).

#### A.5 Detailed number of clinical questions and systematic reviews for the temporal evolution of the certainty of evidence (manuscript Figure 5)

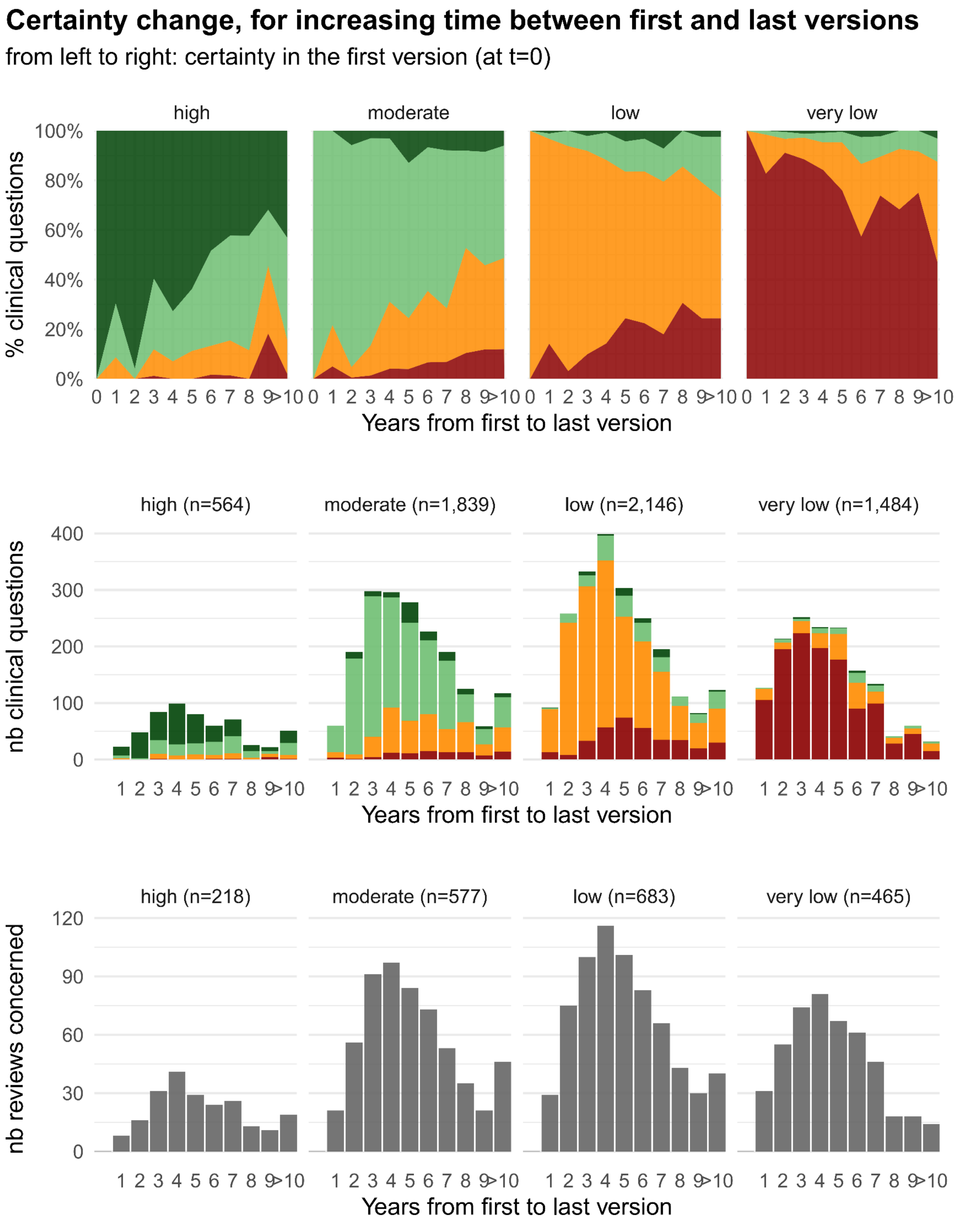


Figure S9. Top panels: Evolution of the certainty of evidence, with increasing interval between the first version (t=0) and the most recent version (x axis). Results are displayed separately for each level of certainty in the first version Middle panels: absolute number of PICO questions concerned, for each year. Bottom panels: absolute number of systematic reviews concerned, for each year.

#### A.6 Effect of additional evidence and time before update on the certainty of evidence

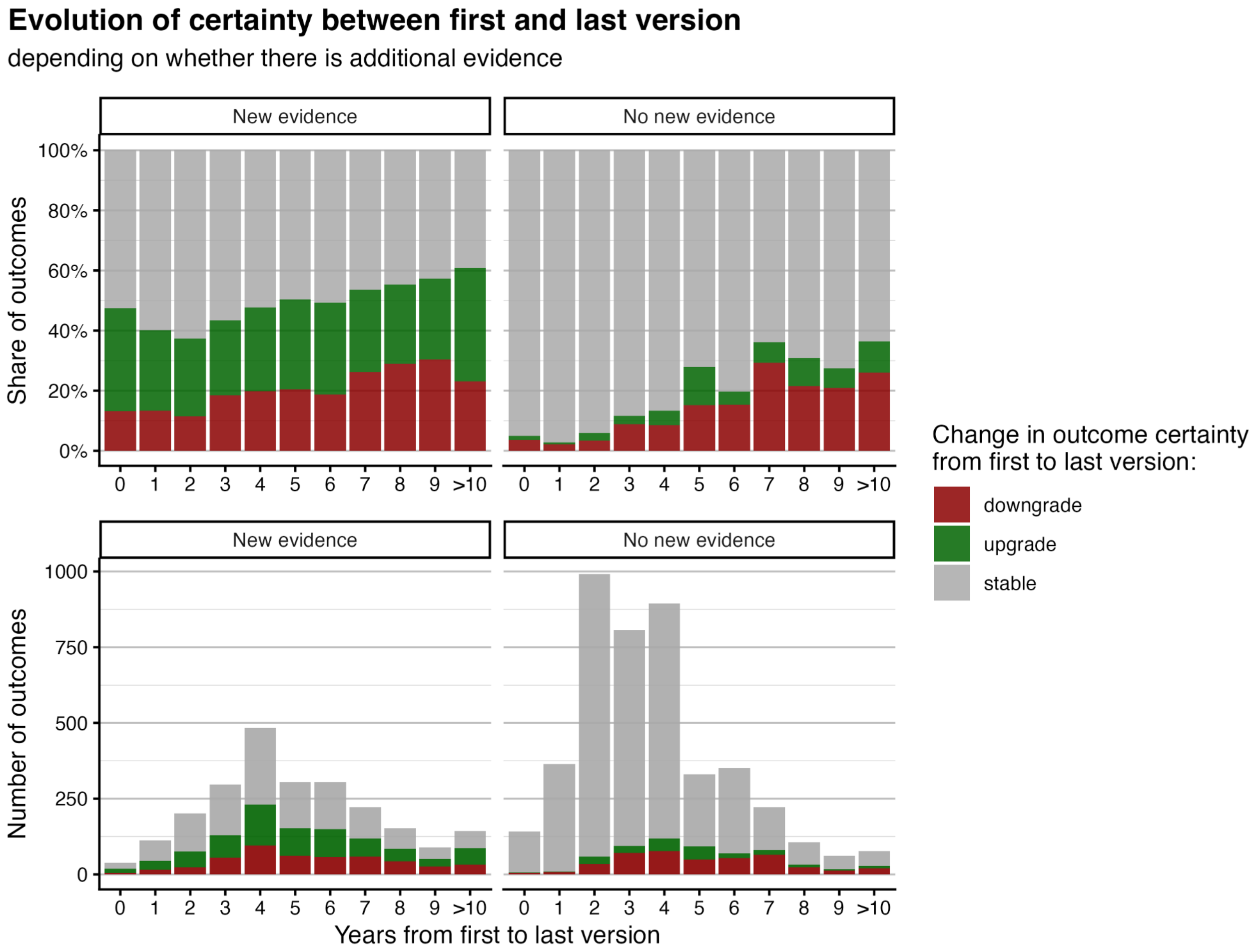


Figure S10: Effect on whether the certainty of evidence is downgraded, upgraded, or stable, depending on the time before the update, and whether additional evidence (number of studies and participants) is included (left and right panes).

### B LLM-assisted data extraction and matching

#### B.1 Structure of the data on the website

Each review has a dedicated version history page (see Figure S5), with a table gathering data of current and past versions of the review. Starting 2010, more and more reviews versions include a Summary of Findings table (example of such a table in Figure S6, alongside the elements we extracted).

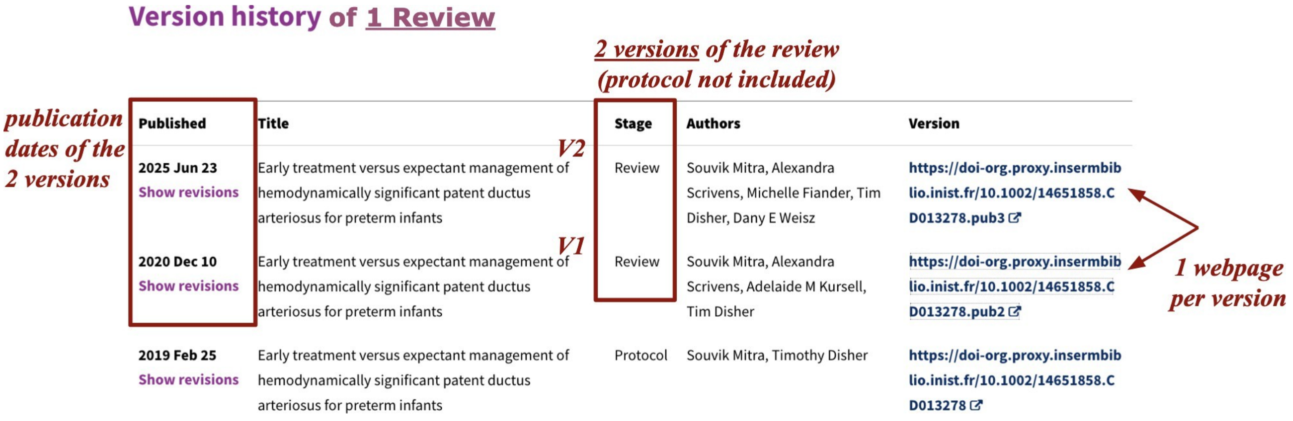


Figure S11: Structure of Cochrane Data: Reviews, Versions

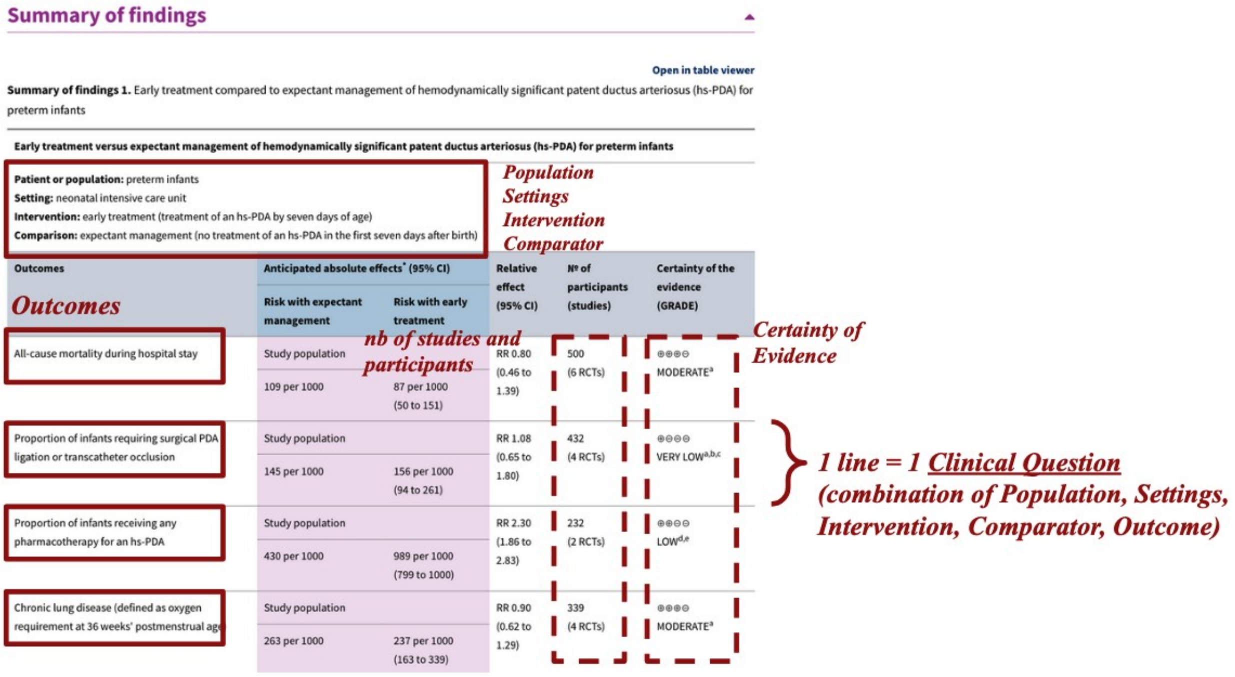


Figure S12: Example of a Summary of Findings table. Each line is defined by a combination of Population, Settings, Intervention, Comparison and Outcome, defining a unique PICO question. We extracted these items, alongside the reported certainty of evidence (not available / very low / low / moderate / high). We also extracted the number of participants and studies, and whether they were randomized or non randomized.

#### B.2 LLM extraction of PICO items and certainty of evidence

Table S1: Extraction performance of the LLM for the 2,660 PICO clinical questions (from 245 versions of 92 distinct reviews) manually verified by human reviewers, for the different extracted items. See text for definition of minor and major error.

| extracted element | correct | minor error | missing line | major error |
|---|---|---|---|---|
| **population** | 98.9% (2631) | 0.0% (0) | 1.1% (29) | 0.0% (0) |
| **settings** | 98.9% (2631) | 0.0% (0) | 1.1% (29) | 0.0% (0) |
| **intervention** | 98.9% (2631) | 0.0% (0) | 1.1% (29) | 0.0% (0) |
| **comparison** | 98.9% (2631) | 0.0% (0) | 1.1% (29) | 0.0% (0) |
| **outcome** | 97.7% (2600) | 1.0% (27) | 1.1% (29) | 0.2% (4) |
| **outcome supp info** | 97.3% (2588) | 0.4% (10) | 1.1% (29) | 1.2% (33) |
| **certainty of evidence** | 98.8% (2627) | 0.1% (3) | 1.1% (29) | 0.0% (1) |
| **nb of studies** | 98.8% (2628) | 0.0% (0) | 1.1% (29) | 0.1% (3) |
| **nb of participants** | 98.4% (2618) | 0.1% (3) | 1.1% (29) | 0.4% (10) |

First, both reviewers independently reviewed a common subset of 25 review versions (309 clinical questions in total), and checked the extracted Summary of Findings tables (see Figure S5). Within this sample, they found 2 missing lines, and 3 major errors concerning the extraction of outcomes; depending on the items, this corresponds to a success rate between 98.4% and 99.4%. There was only 1 disagreement between the reviewers verification (1 extraction error spotted by 1 reviewer and not the other), for the item "number of participants": the inter-reviewer raw agreement was 99.5% for this iem, and 100% for the other items. Consequently, for the remaining 220 review versions, they continued the verification without independent double review.

In total, the verification process concerned 245 versions from 92 distinct reviews, representing 2,660 clinical questions. The distribution of correct extractions and errors is presented in Table S1.

Missing lines accounted for 1.1% (29) of PICO clinical questions. They appear to mainly occur in very long tables, for which the LLM seems to truncate the result. We think it is unlikely to bias our extracted dataset, and argue that this omission behaves similarly to sampling ~ 99% of Cochrane Summary of Findings table content.

Major error rate was 0.0% for items population, setting, intervention, comparison, and certainty of evidence; 0.1% for the number of studies; and 0.4% for the number of participants. More major errors occurred for the outcome item (1.2%), but of the 33 major errors observed, 27 originated from a single review (CD013756). Excluding this single review, the remaining 5 errors represent an error rate of 0.2%.

#### B.3 LLM extraction of search dates

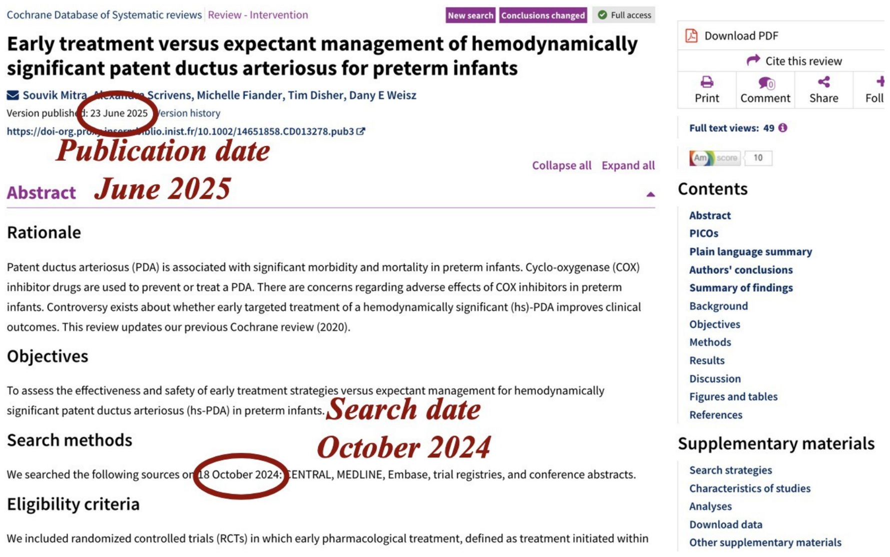


Figure S13: Official publication date, and search date in the Search methods section of the abstract, exemplified for 1 particular review version on Cochrane website.

We manually extracted 600 literature search dates, to understand the logic of the search date reporting in Cochrane reviews. It appeared that:

1. the relevant information is almost always present in the abstract, meaning that consulting the full text is usually unnecessary;
2. most often, the abstract reports a specific search date (e.g. "searched on May 2019"), however in some cases no explicit search date is provided but an inclusion criteria is given instead (e.g. "from Sept 1999 to Apr 2014"); in this case, we focus on the upper bound of the inclusion timeframe.
3. sometimes multiple databases are searched with different cut-off dates; in these cases we decided to only keep the most recent one;
4. abstracts sometimes mention dates corresponding to previous versions of the review, followed by a description of a new search undertaken for the current update. Such earlier dates refereing to previous versions must be ignored.

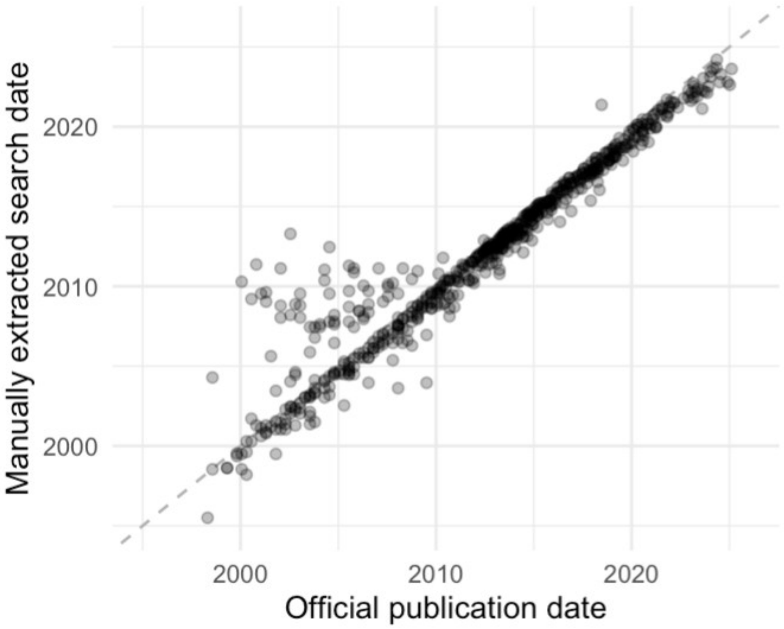


Figure S14: Comparison of manually extracted literature search dates with official publication dates for 600 systematic reviews. In most cases, the search date precedes the official publication by a few months. Larger discrepancies also appear, particularly in the cluster of points observed between 2000 and 2010, where some search dates even fall years after the publication date. These inconsistencies arise from reviews whose updates did not generate a new DOI, leading to silent webpage modifications. This explains why the official publication date could not be used systematically.

From these rules, we optimized a prompt on these 600 observations (training set), to automatically extract with a LLM (GPT-o4-mini-high), from abstract Search Methods section, the search month and year, or, when absent, the upper bound of the inclusion period (final prompt available in Section C.2). If the LLM was unable to extract a date, we used the "New search" events date; if no such events was available, we used the publication date. We then tested our method on 600 additional manually-extracted dates (testing set), for which we presen the results below.

Out of the 14,928 review versions sent to the LLM, 91% (13,592) had a successfully extracted search date; for the remaining 9% unsuccessful ones, it was estimated with the event ”New search” date if available (2% of cases), or with official publication date otherwise (7%). In 84% of the 600 versions tested, our search date estimation matched exactly (month–year) the one manually extracted by the two medical students. In 3.5% and 1.8% of cases, respectively, the estimated date deviated by more than 6 months and more than 1 year. Consequently, we judge that our extraction provided a reliable approximation of the true search date. The raw comparison of our LLM-assisted extraction to the 600 manually-extracted dates is also shown on Figure S7.

Table S2: Performance of our LLM-assisted extraction of the Search dates. Benchmark: 600 manually-extracted search dates from abstracts. We also show the performance for the naive methods only using available metadata (publication date and events date), to show the improvement.

| Estimation method | Pearson $R^2$ | % off by >6 months | % off by >1 year | months difference 2.5 –97.5% quintiles |
|---|---|---|---|---|
| LLM-assisted | 0.998 | 3.5% | 1.8% | -7 – 1 |
| Events + Publication date | 0.981 | 49% | 21% | -28 – 6 |
| Publication date only | 0.917 | 62% | 32% | -28 – 70 |

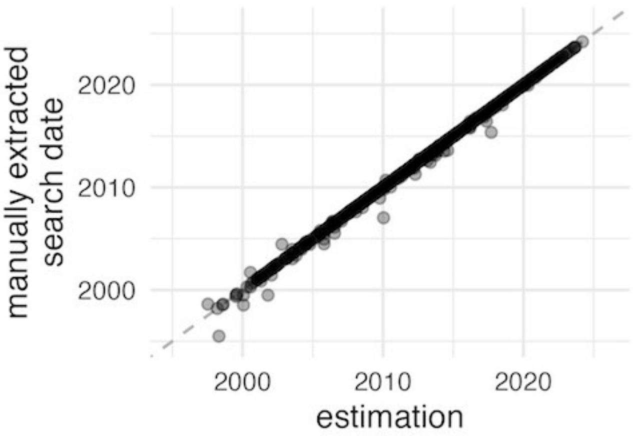


Figure S15: comparison of our LLM-assisted extraction to 600 manually-extracted dates.

#### B.4 LLM matching of clinical questions

Table S3: Top: reviewers judgement (after consensus) with the LLM decision concerning the matching of the PICOs. Bottom: inter-reviewer agreement.

| | population | setting | intervention | comparison | outcome | global |
|---|---|---|---|---|---|---|
| | | | **reviewers judgement of the LLM-matching:** | | | |
| agree - completly | 96.8% | 86.5% | 96.0% | 95.1% | 86.8% | **81.8%** |
| agree - mostly | 1.8% | 12.3% | 3.0% | 3.9% | 11.8% | **16.3%** |
| disagree - mostly | 1.0% | 0.5% | 0.5% | 0.5% | 1.0% | **1.3%** |
| disagree - compltely | 0.4% | 0.6% | 0.4% | 0.4% | 0.4% | **0.6%** |
| | | | **inter-reviewer agreement**: | | | |
| raw | 98% | 94% | 97% | 96% | 94% | **91%** |
| kappa | 95% | 90% | 94% | 92% | 90% | **86%** |

### C Prompts used with the Large Language Models

#### C.1 Prompt for Summary of Findings table extraction

I have the following Cochrane Summary of Findings section (as HTML), containing one or multiple tables. Each table presents multiple outcomes with various details. Also, do not extract any data from the comments column.

For each outcome, extract the following information and return the data as a JSON object that exactly conforms to the provided JSON schema. The JSON object must have a key '"results"' whose value is an array of outcome objects.
Each outcome object must include the following keys:

- **SOF_title:** the table title (usually the first line) describing what is evaluated.
- **patient_population:** the population (and possibly subpopulations) of interest.
- **settings:** e.g., inpatient hospital, primary care in Europe, etc.
- **intervention:** the experimental intervention.
- **comparison:** the comparator intervention (including no specific intervention).
- **outcome:** the outcome measured.
- **outcome_additional_info:** any additional information about the outcome (e.g., scale, follow-up, etc.). If you have nothing to write, write 'NA'.

- **certainty_of_evidence:** the Certainty of Evidence (without symbols such as + or -). Use one of the following values: '"high"', '"moderate"', '"low"', '"very low"', '"NA"', or '"unable to attribute"'.

- **number_of_participants:** the number of participants as an integer.
- **nb_total_studies:** the total number of studies as an integer (sometimes the sum of different types of studies).

- **remark_gpt:** if you have any doubts or difficulties extracting any item, provide a remark here. Also, if the table relates to a network meta-analysis (e.g., there is a Ranking SUCRA), write "network meta-analysis". If you have nothing to say, write ‘"NA"‘.

**Return only a valid JSON object matching this structure with a top-level key ‘"results"‘. Do not include any additional text, Markdown formatting, or code blocks.**

Below is the combined HTML content for the tables:

[here is included the HTML code of the Summary of Findings table, scrapped from the Cochrane website]

#### C.2 Prompt for review Search date extraction

You are a specialised assistant that extracts two dates from the Search Methods section of Cochrane review abstracts. For each record you receive (see the embedded JSON array) return exactly:

• `cochrane_ID`: string (copied verbatim)
• `search_year`: 4-digit integer, or null if no valid search-date year is found
• `search_month`: three-letter English month abbreviation (`Jan`, `Feb`, `Mar`, `Apr`, `May`, `Jun`, `Jul`, `Aug`, `Sep`, `Oct`, `Nov`, `Dec`) or `NA` if the month is absent / not inferable
• `inclusion_year`: 4-digit integer, or null if no valid inclusion-end year is found
• `inclusion_month`: three-letter English month abbreviation or `NA` if the month is absent / not inferable

### Rules
1. Infer only from the provided `Text` field.
2. Consider only years between 1995 and 2025 inclusive.
3. Build the list of **search-date** candidates by finding any date that clearly marks the **current search run** (e.g. "searched on 12 March 2021", "last searched October 2001").
4. Build the list of **inclusion-end** candidates by finding the **upper bound** of any inclusion-interval expression (e.g. "studies published between January 2000 and May 2020", "eligibility up to Feb 2022"). *Ignore* the lower bound of ranges ("from 1966").
5. For both dates (search and inclusion) exclude explicit dates labelled as belonging to a previous version of the review (e.g. "previous search", "earlier version"). We focus on the current version / update.
6. For each set of candidates (search and inclusion), if multiple dates remain:
   a. Pick the date that appears **most frequently** in the text (case-insensitive exact matches of the year or year-month string).
   b. If two or more dates tie on frequency, choose the one with the **latest** calendar value (year → month → day).

7. If a chosen date includes only a year, set its corresponding month field to

`"NA"`.
8. If no valid candidate date exists for a field, set its year to `null` and its month to `"NA"`.
9. Respond *only* with JSON that conforms to the schema—no extra keys, comments, or free text.

Return an object with a single key `"results"`, whose value is an array of objects each containing the five fields above.

### Examples

#### Example 1
##### Input
```json
[
  {
    "cochrane_ID": "CD002290",
    "Text": "The Cochrane Pregnancy and Childbirth Group trials register and the Cochrane Controlled Trials Register were searched. Date of last search: October 2001."
  }
]
```

##### Output
```json
{
  "results": [
    {
      "cochrane_ID": "CD002290",
      "search_year": 2001,
      "search_month": "Oct",
      "inclusion_year": null,
      "inclusion_month": "NA"
```

    }
  ]
}
```

## Example 2

### Input
```json
[
  {
    "cochrane_ID": "CD003966",
    "Text": "We searched the Cochrane Central Register of Controlled Trials (CENTRAL in The Cochrane Library) MEDLINE (from 1966) and EMBASE (from 1988) without language restriction."
  }
]
```

### Output
```json
{
  "results": [
    {
      "cochrane_ID": "CD003966",
      "search_year": null,
      "search_month": "NA",
      "inclusion_year": null,
      "inclusion_month": "NA"
    }
  ]
}
```

### Explanation
```

The only dates present refer to the beginning of the interval, but there is no mention of up to which date the search was performed.

#### Example 3

##### Input
```json
[
  {
    "cochrane_ID": "CD003004",
    "Text": "We searched the Cochrane Central Register of Controlled Trials (CENTRAL) (The Cochrane Library 2012, Issue 9); MEDLINE (1966 to October 2012); and EMBASE (1980 to October 2012). This is an updated version of a review published in 2004. The original search was performed in October 2003. "
  }
]
```

##### Output
```json
{
  "results": [
    {
      "cochrane_ID": "CD003004",
      "search_year": null,
      "search_month": "NA",
      "inclusion_year": 2012,
      "inclusion_month": "Oct"
    }
  ]
}
```

##### Explanation

The years 2004 and 2003 refer to earlier versions. The current searches were run up to October 2012, which are the upper bound of the inclusion interval. There is no mention of the date when the search was performed.

### Record from which you extract data:

[HERE IS INSERTED THE DATA]

#### C.3 Prompt for matching similar research questions across review versions

#### Context

You have a JSON array of Cochrane “Summary of Findings” exported from multiple versions of the same systematic review. Each object has:
- a unique `"id"`
- a `"version_stage"` (e.g. `"V1"`, `"V2"`, ...)
- seven key fields that together define a research question R:
  1. `"SOF_title"`
  2. `"patient_population"`
  3. `"settings"`
  4. `"intervention"`
  5. `"comparison"`
  6. `"outcome"`
  7. `"outcome_additional_info"`

Inside each version, no two objects share the same research question R. However across versions, the same question R can be addressed. We need to identify which Rs are the same **across different versions only**. However across versions, a similar R is probably be re-worded; it probably won't be a perfect word-for-word match.

#### Task

##### Goal

Cluster objects whose research questions **R** are semantically equivalent **across different versions only**.

##### Hard constraints (must be satisfied before you return the answer)

1. **Cross-version only**
   - Only link questions R if their `version_stage` values differ.

2. **Uniqueness within version**
   - Within any single `version_stage`, each non-null `match_id` should appear **at most once**.

3. **Minimum evidence**
   - Any non-null `match_id` must appear in **at least two different** `version_stage`s.
   - After applying rules 1–2, if a `match_id` is used only once, change it to `null`.

##### Quick example

**Input**
```json
[
 {
```

"id": 13421,
"version_stage": "V3",
"SOF_title": "Communication skills training compared with no communication skills training for improving healthcare professionals (HCP) communication with cancer patients",
"patient_population": "healthcare professionals working with patients with cancer",
"settings": "outpatient or primary care",
"intervention": "A communications skills training program",
"comparison": "No communication skill training",
"outcome": "HCP showed 'empathy'",
"outcome_additional_info": "NA"
},
{
"id": 13422,
"version_stage": "V3",
"SOF_title": "Communication skills training compared with no communication skills training for improving healthcare professionals (HCP) communication with cancer patients",
"patient_population": "healthcare professionals working with patients with cancer",
"settings": "outpatient or primary care",
"intervention": "A communications skills training program",
"comparison": "No communication skill training",
"outcome": "HCP used 'open questions'",
"outcome_additional_info": "NA"
},
{
"id": 13424,
"version_stage": "V3",
"SOF_title": "Communication skills training compared with no communication skills training for improving healthcare professionals (HCP) communication with cancer patients",
"patient_population": "healthcare professionals working with patients with cancer",
"settings": "outpatient or primary care",
"intervention": "A communications skills training program",
"comparison": "No communication skill training",
"outcome": "Patient satisfaction with communication",
"outcome_additional_info": "NA"
},
{
"id": 20652,
"version_stage": "V4",
"SOF_title": "CST compared to control for healthcare professionals working with people who have cancer",
"patient_population": "Healthcare professionals working with people who have cancer",
"settings": "NA",
"intervention": "Communication skills training",
"comparison": "No communication skills training",
"outcome": "Showed empathy",
"outcome_additional_info": "NA"
},

{
"id": 20649,

"version_stage": "V4",
"SOF_title": "CST compared to control for healthcare professionals working with people who have cancer",
"patient_population": "Healthcare professionals working with people who have cancer",
"settings": "NA",
"intervention": "Communication skills training",
"comparison": "No communication skills training",
"outcome": "Used open questions",
"outcome_additional_info": "NA"
},
{
"id": 20650,
"version_stage": "V4",
"SOF_title": "CST compared to control for healthcare professionals working with people who have cancer",
"patient_population": "Healthcare professionals working with people who have cancer",
"settings": "NA",
"intervention": "Communication skills training",
"comparison": "No communication skills training",
"outcome": "Elicited concerns",
"outcome_additional_info": "NA"
}
]
```

**Output**
```json
[
{ "id": 13421, "match_id": 1 },
{ "id": 20652, "match_id": 1 },
{ "id": 13422, "match_id": 2 },
{ "id": 20649, "match_id": 2 },
{ "id": 13424, "match_id": null },
{ "id": 20650, "match_id": null }
]
```

### Output schema

Return only the JSON array above. Each element must be

{ "id": <integer>, "match_id": <integer|null> }

## Data to process:

[HERE IS INSERTED THE DATA]
```
