## SupplementaryMethods for "Amount and certainty of evidence in Cochrane systematic reviews of interventions: a large-scale meta-research study"

### Supplementary Methods: use of LLM for data extraction and processing

#### Data retrieved through web scraping

For each review version, the following data were automatically extracted during our web scraping:

- metadata: identifier, version, title, URL, official publication date, and dates of revision events (e.g., "new search performed", "conclusion changed");
- "Search Methods" text from the abstracts: Cochrane abstracts are always structured with the same subsections, which include a "Search Methods" section. We retrieved this section during our web scraping. This was then processed by a large language model (LLM) to extract the literature search dates.

**Data extraction assisted by a large language model (LLM)**

We extracted the following information, along with a record of its source, using an LLM: 1) the search date of each version of each systematic review, 2) information reported in the “Summary of findings” tables (population, intervention, comparison, outcome [PICO] questions; certainty of evidence; number of studies; and number of participants). In addition, when previous review versions reporting a “Summary of findings” table were identified, we used an LLM to match the PICO questions between the most recent update and the initial version with a “Summary of findings” table. The specific procedures for each task are detailed below.

<u>Search date of the systematic reviews</u>

The publication date of the reviews can differ substantially from the search date (Supplementary Figures S13 and S14). We extracted the search date from the "Search Methods" section of each abstract, with GPT-o4-mini-high . If the LLM extraction returned an empty value (for instance, because a search date was not mentioned in the abstract), we relied on the review version metadata (i.e., "new search" date) when available; as a last resort, we relied by default on the official publication date as an approximation.

We optimized prompts to automatically extract the search month and year with GPT-o4-mini-high on a training set of 600 review "Search Methods" sections manually extracted by two specifically trained medical students, each record being extracted by a single reviewer. We used few-shot prompting with 3 representative examples provided to guide the model (see Supplementary Section B.3, C.2).

This optimized prompt was then validated on an independent testing set of 600 additional search dates, manually extracted by the two reviewers with each record extracted by a single reviewer. Overall, the LLM-assisted extraction exactly matched the manually extracted month–year in 84% of the 600 cases; 98% were within ±1 year, and errors ranged from −3.0 to +1.7 years.

<u>"Summary of findings" table: PICO question, and quantity and certainty of evidence for each outcome</u>

In Cochrane reviews, a "Summary of findings" table is dedicated to a single combination of population-intervention-comparison (PIC); within the table, each line reports the number of studies and certainty of evidence for a single outcome (meaning that each line of the table represents a unique combination of the PICO elements that defines a clinical question). Several "Summary of findings" tables can be reported within a single review. When a "Summary of findings" table was reported, we extracted the following information (see Supplementary Figure S12 for the structure of a "Summary of findings" table on the Cochrane website):

- PICO questions: population, settings, intervention, comparison, and outcome.
- The number of studies and participants included for the evaluation of each PICO reported in each "Summary of findings" table.
- Whether the number of studies refers to randomized controlled trials (RCTs) or non-randomized studies; this information could also be extracted from the abstracts section "Selection Criteria" if needed.
- The certainty of the evidence for each outcome reported in each "Summary of findings" table. The certainty of evidence was rated as high, moderate, low, very low, or unassessed when insufficient evidence was found.

Because "Summary of findings" tables vary slightly in layout and structure, a deterministic scraping extraction was not appropriate. Consequently, we used an LLM to extract this information.

We provided the raw HTML code displaying the "Summary of findings" tables to an LLM (GPT-o3-mini-high, OpenAI's most recent mini-reasoning model available in April 2025). We developed prompts that were iteratively optimized on 15 reviews (the final prompt is available in Supplementary Section C.1). As the "Summary of findings" tables comprise highly structured tabular data, we used zero-shot prompting (i.e., no example given in the prompt).

The accuracy of the LLM data extraction was assessed in a random sample of 245 review versions (92 unique reviews, containing 2,660 PICO questions) manually checked by two specifically trained medical students.

They classified the extraction generated by the language model into four categories:

- **Correct**: the extracted text was identical to the text reported in the Cochrane "Summary of findings" table.

- **Minor error**: the extracted text differed slightly but without altering the interpretation. For example, for review CD006715, the outcome was “Pulmonary complications (0 to 30 days): Respiratory depression” on the Cochrane website, whereas the LLM extracted “Respiratory depression: (0 to 30 days)”.
- **Missing line**: an entire line (PICO question) of the “Summary of findings” table was omitted, with all its elements. This typically occurred in reviews with particularly long tables, or with many tables.
- **Major error**: the extracted text differed from the original in a way that affected interpretation.

As quality control, the two reviewers independently checked in duplicate a subset of 309 PICO questions retrieved from 25 reviews. The raw agreement between the two reviewers was high: 99% to 100% depending on the elements (we did not use Cohen's kappa to judge agreement because the error rates were very low [24]; details in Supplementary Section B.2). Considering this high agreement, the remaining 2,351 PICO questions were divided between the two reviewers for single assessment.

Overall, among 2,660 PICO questions that were manually verified, 98% were judged to be correctly extracted, 1% were missed, and 1% were extracted but with major errors (see Supplementary Section B.2). Most major errors came from a single review (CD013756); when this single review was excluded, the major error rate dropped to 0.2%.

<u>Automatic matching of similar PICO questions with previous versions of the review</u>

To assess the evolution of evidence, we matched similar PICO questions present in the most recent update of a review and its initial versions that reported a “Summary of findings” table, when available. Because the wording of the PICO elements for the same clinical question may vary between versions (e.g., “participants with unexplained subfertility” vs “people with unexplained subfertility” for the population in review CD001838), we used LLM-assisted matching (gpt-o4-mini-high).

We provided the LLM with the review version identifiers and the PICO elements previously extracted from the “Summary of findings” tables into an Excel sheet. The associated certainty of evidence was not provided to the LLM, to avoid influencing the model’s decision.

We optimized a prompt on 100 reviews. If a PICO question in one review version had no corresponding question in other versions, it was left unmatched. The final prompt is available in Supplementary Section C.3 (few-shot prompting, with one representative example).

For quality control, two reviewers independently assessed whether the matches identified by the LLM were consistent on a random sample of 181 reviews (425 distinct versions) addressing 4,853 PICO questions. The reviewers rated the level of agreement in four categories: complete agreement, mostly agree, mostly disagree, and complete disagreement. Any discrepancies were resolved by a third reviewer. The PICO question was classified as similar between review versions only if all elements of the PICO (population-intervention-comparison-outcome) were similar.

Overall, the human reviewers agreed with the LLM’s decisions in 98% of cases. The inter-reviewer raw agreement was 99%. The more granular assessment of the similarity between matches was as follows: complete agreement (82%), mostly agree (16%), mostly disagree (1%), and complete disagreement (0.6%). Inter-reviewer reliability was high (Cohen’s $\kappa$=86%). More details for each PICO element are available in Supplementary Section B.4.
