## Supplementary material for "Amount and certainty of evidence in Cochrane systematic reviews of interventions: a large-scale meta-research study": Checklist_Meta_epidemiological_Study

### Checklist for Reporting Meta-Epidemiological Methodology Research

*Adapted from Murad MH, Wang Z. Guidelines for reporting meta-epidemiological methodology research.* ***Evid Based Med***. 2017;22(4):139–142.

**Manuscript Title:** Amount and certainty of evidence in Cochrane systematic reviews of interventions: a large-scale meta-research study

**Authors:** Thomas Starck, Philippe Ravaud, Isabelle Boutron

| Section/ Topic | Checklist Item | Reported | Location in Manuscript | Comments |
|---|---|---|---|---|
| **Title** | Identify the report as a meta-epidemiologic study | Yes | Title page | Title identifies study as "large-scale meta-research study" (acceptable synonym for meta-epidemiological) |
| **Abstract Structured summary** | Provide a structured summary that includes the background of the topic, goal of the study, data sources, method of data selection, appraisal and synthesis methods, results, limitations, conclusions and implications of key findings | Yes | Abstract | Structured abstract present with all required elements<br>- background: OK<br>- goal of the study: OK (objectives)<br>- data sources: OK<br>- method of data selection: OK (eligibility criteria + data extraction)<br>- appraisal and synthesis methods: OK (analysis)<br>- results: OK<br>- limitations: OK<br>- conclusions: OK<br>- implications of key findings: OK |
| **Introduction Rationale** | Describe the rationale for the meta-epidemiological study in the context of what is already known | Yes | Introduction | |
| **Introduction Objectives** | Provide an explicit statement of the goal of the meta-epidemiological study and the hypothesis being empirically tested | Partial | Introduction (final paragraph) | Our objective is to describe the evolution of the amount and certainty of evidence; we do not formally test an hypothesis |
| **Methods Protocol** | Indicate if a protocol exists, if and where it can be accessed. Registration of a protocol is not mandatory | Yes | NA (no protocol registered) | Protocol registration not mandatory for meta-epidemiological studies; we de not test hypotheses or perform formal statistical test, so our design is not subject to p hacking |
| **Methods Eligibility** | Specify study characteristics used as | Yes | Methods - Eligibility | |

| | | | | |
|---|---|---|---|---|
| **criteria** | criteria for eligibility with a rationale | | Criteria | |
| **Methods Information sources** | Describe all information sources (databases with dates of coverage) and search date | Yes | Methods - Search strategy | Cochrane Database search date (April 8, 2025) reported |
| **Methods Search** | Present full electronic search strategy for at least one database, such that it could be repeated | Not Applicable | | Search equations replaced by an exhaustive analysis of the entire Cochrane Database of Systematic Reviews. |
| **Methods Study selection** | Describe the process for selecting studies for inclusion (how many reviewers, reviewing in duplicate or single) | Not Applicable | | Inclusion of systematic reviews was automatically detected based on the presence / absence of Summary of Findings |
| **Methods Data collection process** | Describe method of data extraction from reports (piloted forms, independently, in duplicate) | Yes | Methods - Data extraction assisted by LLM and Supplementary Materials | LLM-assisted extraction with human quality control on a sample; inter-reviewer agreement reported |
| **Methods Data items** | List and define all variables for which data were sought and any assumptions and imputations made | Yes | Methods - Data retrieved / Data extraction assisted by a LLM | |
| **Methods Risk of bias in individual studies** | If risk of bias assessment of individual studies was relevant to the analysis, describe the items used | Not Applicable | | Not applicable to this meta-epidemiological study design (reporting content of systematic reviews) |
| **Methods Summary measures** | State the principal summary measures and explain its meaning and direction to readers | Yes | | We only use standard descriptive statistics: proportions, median, Q1-Q3. No explanation needed. |
| **Methods Synthesis of results** | Describe the statistical or descriptive methods of synthesis. Describe methods of additional analyses if done | Yes | | Same as above |
| **Results Study selection** | Give numbers of studies assessed and included, with reasons for exclusions, ideally with a flow diagram. Present inter-reviewer agreement | Yes | Figure 1 for flow chart Methods for inter-reviewer agreement: (kappa, raw agreement) | |
| **Results Study characteristics** | For each study, present characteristics for which data were extracted. Clinical characteristics may not always be relevant | Yes | Results - Identification and characteristics of Cochrane Reviews | |
| **Results Risk of bias within studies** | If risk of bias assessment was used, report indicators of each study | NA | | Not applicable to this meta-epidemiological study design (reporting content of systematic reviews) |
| **Results Results of** | Present data elements used in the meta-epidemiological analysis | Yes | Results; Tables and Figures | Certainty of Evidence, number of studies, participants, presented per PICO |

| | | | | |
|---|---|---|---|---|
| **individual studies** | from each study | | | |
| **Results Synthesis of results** | Present results of statistical analysis done, including measures of precision and consistency | Yes | Results; Figures 2-5; Table 1 | Results presented with medians and quartiles in text |
| **Results Additional analysis** | Give results of additional analyses, if done (e.g., sensitivity or subgroup analyses) | Yes | Results - Certainty of evidence Sensitivity analysis; Supplementary Figure S10 | Sensitivity analysis for PICOs with no new evidence |
| **Discussion Summary of evidence** | Summarize the main findings and compare with existing knowledge. Investigators should describe their certainty in the results to readers | Yes | Discussion - Statement of principal findings and comparison with literature | |
| **Discussion Limitations** | Discuss limitations at research methodology level (e.g., likelihood of reporting or publication bias) | Yes | Discussion - Strengths and weaknesses of the study | |
| **Discussion Conclusions** | Provide general interpretation of the results and implications for future research. Provide any plausible impact on clinical practice | Yes | Discussion - Meaning of the study; Conclusion | |
| **Funding** | Describe sources of funding for the methodology research and role of funders | Yes | Statements - Role of funding source | |
